# Metabolomics of IgE-Mediated Food Allergy and Oral Immunotherapy Outcomes based on Metabolomic Profiling

**DOI:** 10.1101/2024.05.31.24308233

**Authors:** Yamini V. Virkud, Jennifer N. Styles, Rachel S. Kelly, Sarita U. Patil, Bert Ruiter, Neal P. Smith, Clary Clish, Craig E. Wheelock, Juan C. Celedón, Augusto A. Litonjua, Supinda Bunyavanich, Scott T. Weiss, Erin S. Baker, Jessica A. Lasky-Su, Wayne G. Shreffler

**Affiliations:** University of North Carolina, Chapel Hill. Department of Pediatrics, Division of Allergy and Immunology, Food Allergy Initiative. Chapel Hill, NC; Massachusetts General Hospital for Children, Food Allergy Center. Department of Pediatrics, Massachusetts General Hospital, Boston, MA; Harvard Medical School, Boston, MA; Channing Division of Network Medicine, Brigham & Women’s Hospital, Boston, MA; Broad Institute, Cambridge, MA; Karolinska University Hospital, Unit of Integrative Metabolomics, Institute of Environmental Medicine, Karolinska Institutet, Stockholm, Sweden; Department of Respiratory Medicine and Allergy. Stockholm, Sweden; Children’s Hospital of Pittsburgh of the University of Pittsburgh Medical Center. Division of Pulmonary Medicine. University of Pittsburgh, Pittsburgh, PA 15260, USA; Icahn School of Medicine at Mount Sinai. Department of Genetics & Genomic Sciences and Department of Pediatrics. New York, NY; University of North Carolina, Chapel Hill. Department of Chemistry. Chapel Hill, NC

**Keywords:** Food Allergy, Immunotherapy, IgE-mediated food allergy, Metabolomics, Secondary bile acids Lithocholic acid, Lithocholate, Eicosanoids, Arachidonic acid, Bile acids, Urocanic acid, Histidines, Sustained Unresponsiveness, Transient Desensitization, Remission

## Abstract

**Background:** The immunometabolic mechanisms underlying variable responses to oral immunotherapy (OIT) in patients with IgE-mediated food allergy are unknown.

**Objective:** To identify novel pathways associated with tolerance in food allergy, we used metabolomic profiling to find pathways important for food allergy in multi-ethnic cohorts and responses to OIT.

**Methods:** Untargeted plasma metabolomics data were generated from the VDAART healthy infant cohort (N=384), a Costa Rican cohort of children with asthma (N=1040), and a peanut OIT trial (N=20) evaluating sustained unresponsiveness (SU, protection that lasts after therapy) versus transient desensitization (TD, protection that ends immediately afterwards). Generalized linear regression modeling and pathway enrichment analysis identified metabolites associated with food allergy and OIT outcomes.

**Results:** Compared with unaffected children, those with food allergy were more likely to have metabolomic profiles with altered histidines and increased bile acids. Eicosanoids (e.g., arachidonic acid derivatives) (q=2.4×10^−20^) and linoleic acid derivatives (q=3.8×10^−5^) pathways decreased over time on OIT. Comparing SU versus TD revealed differing concentrations of bile acids (q=4.1×10^−8^), eicosanoids (q=7.9×10^−7^), and histidine pathways (q=0.015). In particular, the bile acid lithocholate (4.97[1.93,16.14], p=0.0027), the eicosanoid leukotriene B4 (3.21[1.38,8.38], p=0.01), and the histidine metabolite urocanic acid (22.13[3.98,194.67], p=0.0015) were higher in SU.

**Conclusions:** We observed distinct profiles of bile acids, histidines, and eicosanoids that vary among patients with food allergy, over time on OIT and between SU and TD. Participants with SU had higher levels of metabolites such as lithocholate and urocanic acid, which have immunomodulatory roles in key T-cell subsets, suggesting potential mechanisms of tolerance in immunotherapy.

**Key Messages:**

– Compared with unaffected controls, children with food allergy demonstrated higher levels of bile acids and distinct histidine/urocanic acid profiles, suggesting a potential role of these metabolites in food allergy.
– In participants receiving oral immunotherapy for food allergy, those who were able to maintain tolerance-even after stopping therapy-had lower overall levels of bile acid and histidine metabolites, with the exception of lithocholic acid and urocanic acid, two metabolites that have roles in T cell differentiation that may increase the likelihood of remission in immunotherapy.

**Capsule summary:** This is the first study of plasma metabolomic profiles of responses to OIT in individuals with IgE-mediated food allergy. Identification of immunomodulatory metabolites in allergic tolerance may help identify mechanisms of tolerance and guide future therapeutic development.

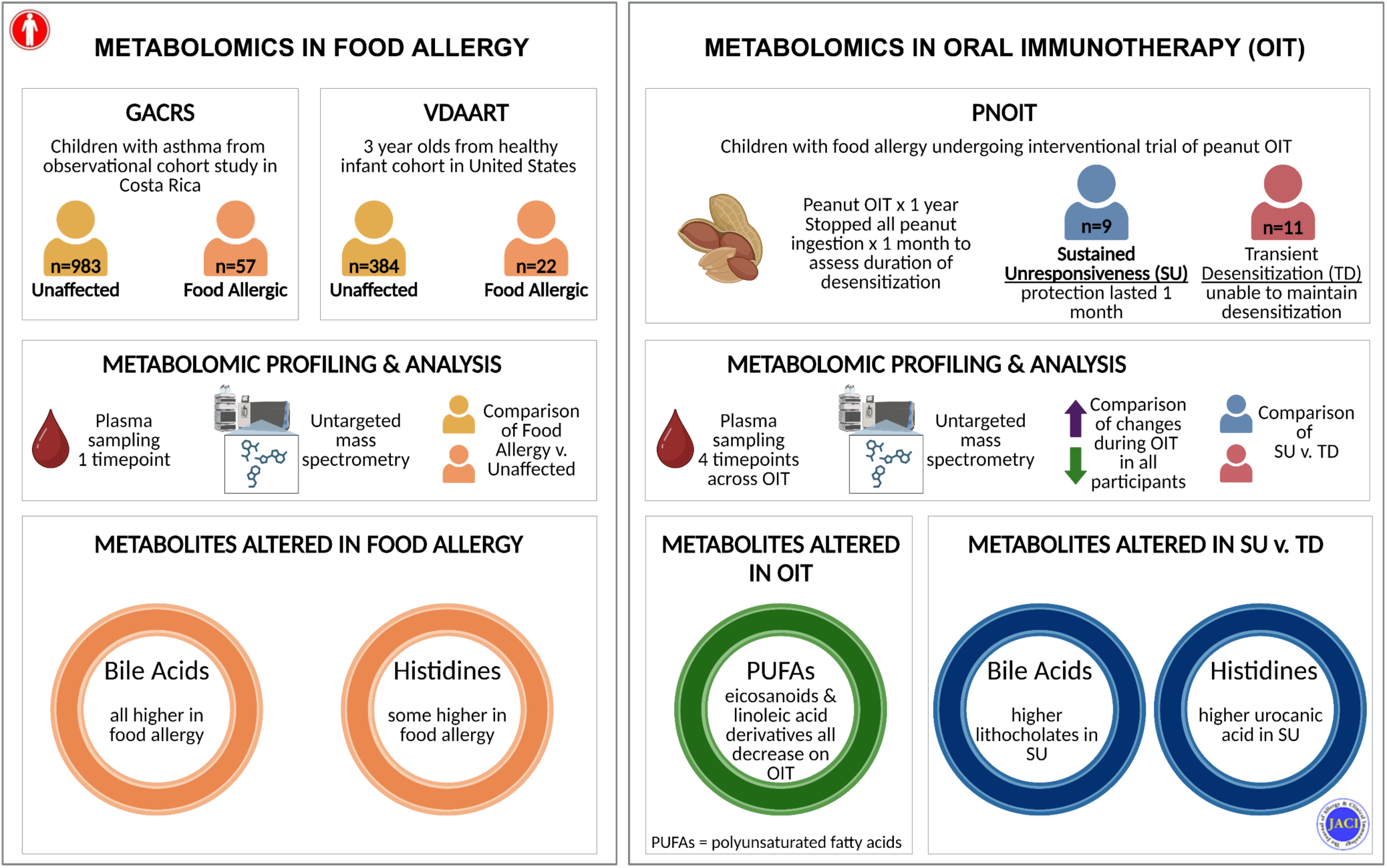

## Introduction

The prevalence of potentially life-threatening immunoglobulin E (IgE)-mediated food allergies is currently estimated at 8-10%, and as the prevalence continues to rise, the need for treatment options becomes more urgent.^1–3^ The development of novel therapies requires a comprehensive understanding of immune tolerance to allergens, including how tolerance differs between individuals with and without food allergies, and how this is modified by treatment.

Although oral immunotherapy (OIT) was approved as the first and only treatment for IgE-mediated peanut allergies in 2020, the effects of such treatment can have limited duration and lead to allergic adverse events.^4^ Most individuals with food allergy (∼50-75%) are capable of achieving transient desensitization, a state where daily OIT protects against accidental exposure to allergen by increasing the eliciting dose for a reaction. While estimates vary by study, only approximately one-third of children demonstrate sustained unresponsiveness (also described as remission), a state of clinical tolerance to large doses of allergen persisting at least weeks after cessation of regular allergen consumption.^5, 6^ Studies of patients on OIT therefore provide a unique insight into the pathophysiology of allergen-specific immunity. Furthermore, while the adaptive immunity underlying food allergies and therapeutic responses have been studied extensively, little is known about the immunometabolism (the interaction between immunity and metabolism) of the modification of tolerance to food allergens.^7–9^

Metabolomics, defined as study of small molecule cellular substrates, intermediates and products of cellular metabolism, can be used to gauge the physiologic state of an organism and can reflect the interaction of the genome, transcriptome, proteome, microbiome, and environmental influences on disease.^10, 11^ Studies support the utility of metabolomic analyses in identifying pathways associated with food allergy, while several investigations have explored the evolution of metabolomic profiles on other types of aeroallergen sublingual and subcutaneous immunotherapy.^9, 12–21^ To date, one study has examined water soluble metabolites in the stool in the setting of food immunotherapy, and none have studied the plasma metabolome in humans.^22^.

Our main objectives were to (1) examine metabolomic profiles of individuals with food allergy in multi-ethnic cohorts (Genetics of Asthma in Costa Rica (GACRS) and the Vitamin D Antenatal Asthma Reduction Trial (VDAART) infant cohort) to identify key pathways of interest, (2) explore how those and other metabolomic pathways evolve with OIT (using the Peanut Oral Immunotherapy trial (PNOIT)), and (3) associate the metabolomic profiles with therapeutic OIT outcomes (**Figure 1**). We used longitudinal sampling to define pathophysiologic differences between those who develop transient desensitization versus sustained unresponsiveness while on OIT, to reveal novel mechanisms in immune tolerance and inform future adjunctive therapies that may shift the response to OIT towards longer-lasting protection.

**Figure 1:**
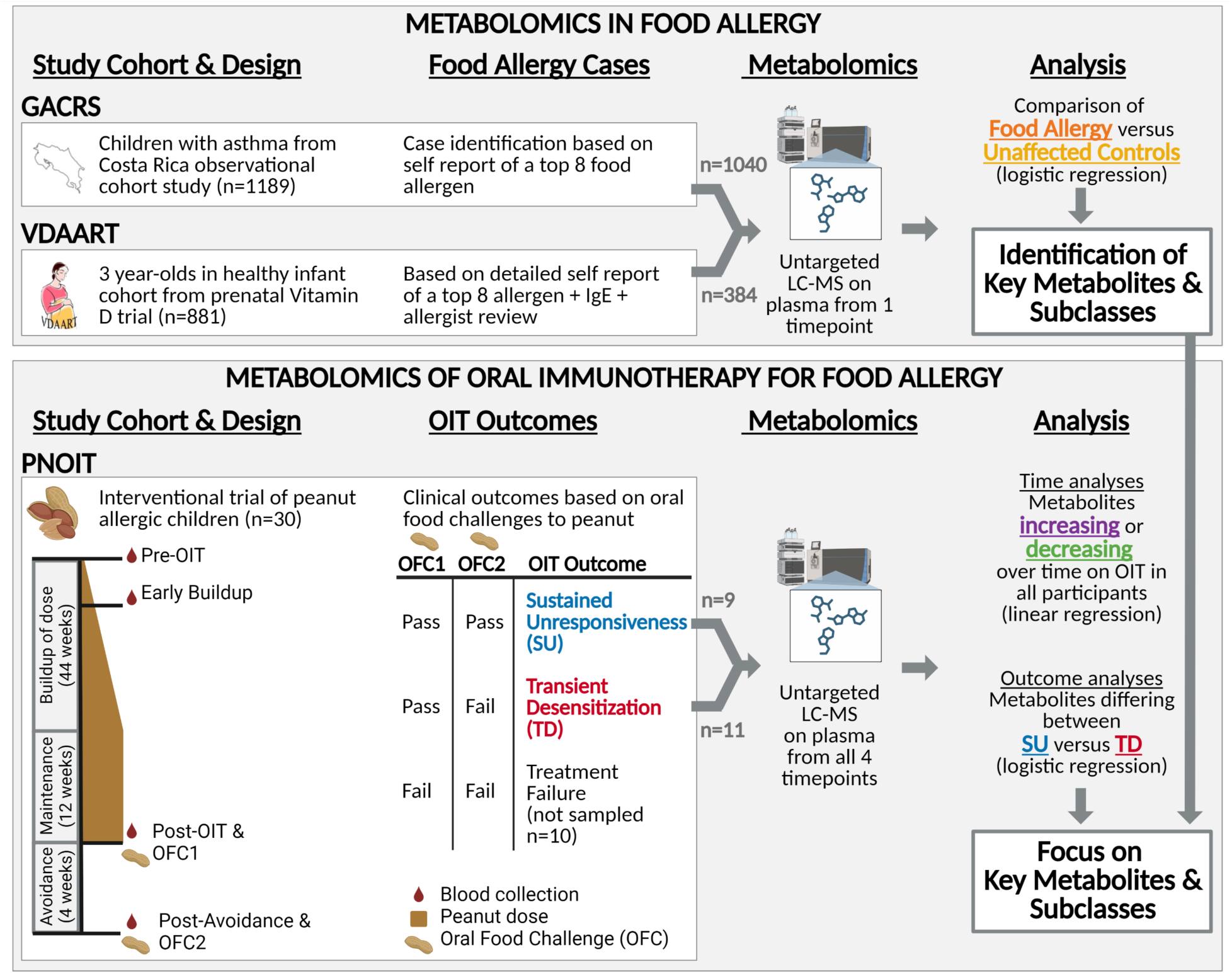
Overview of Cohort Study Design and Analysis in 3 studies. Top: To study the metabolomics of food allergy, 2 cohorts, the Genetics of Asthma in Costa Rica (GACRS) and the Vitamin D Antenatal Asthma Reduction Trial (VDAART) were used. Food allergy status was determined based on available clinical data (self-report for GACRS and detailed food allergy questionnaire for VDAART). Untargeted metabolomic profiles were compared between participants with food allergy and unaffected controls to identify key metabolites and subclasses of interest. Bottom: To study the metabolomics of OIT responses, we used the Peanut Oral Immunotherapy trial (PNOIT), where participants underwent 56 weeks of peanut immunotherapy, followed by 4 weeks of avoidance to determine who could maintain the protection of OIT after stopping (sustained unresponsiveness, SU) versus who lost protection soon after stopping (transient desensitization, TD), as determined by oral food challenges to full servings of peanut. Untargeted metabolomics was performed on the SU and TD groups to determine metabolites that changed over time on OIT versus those that differed between SU and TD groups. Created with BioRender.com.

## METHODS

### Study Populations

Metabolomics data from three populations was compiled to address our objectives: (1) the Genetics of Asthma in Costa Rica Study (GACRS),^23–26^ (2) the Vitamin D Antenatal Asthma Reduction Trial (VDAART),^27^ and (3) the Peanut Oral Immunotherapy (PNOIT) trial.^28^

*GACRS:* Participants in the GACRS are members of a semi-isolated Latin American population from the Central Valley of Costa Rica, which has one of the highest rates of asthma worldwide (24% in children).^23–25^ Children ages 6-14 years were eligible for the study if they had asthma (physician-diagnosed asthma and ≥2 respiratory symptoms or asthma attacks in the prior year) and a high probability of having ≥6 great-grandparents born in the Central Valley of Costa Rica. A total of 1,189 children with asthma and their parents (i.e., 383 parent-child trios) were enrolled. The parents of participating children completed extensive questionnaires, including a slightly modified and translated version of the questionnaire used in the Collaborative Study on the Genetics of Asthma^29^. Metabolomic profiling was available for 1159 participants, and of those food allergy data were available for 1040 participants.

VDAART: The pediatric cohort of VDAART comprises the infants of mothers who were prenatally randomized to Vitamin D supplementation versus placebo in the multi-center trial (ClinicalTrials.gov ID NCT00920621).^27^ All infants had a family history of asthma, eczema or allergic rhinitis in at least one biological parent. Families completed comprehensive questionnaires with detailed phenotyping of allergic diseases, including food allergy. The original study enrolled 881 mothers and their infants. We used metabolomic data (available for 411 participants) from plasma samples collected at age 3, to study the association with food allergy at that age (available for 384 participants).

*PNOIT*: Pediatric participants (aged 7-21) enrolled in an open-label interventional peanut OIT trial, which involved the administration of gradually increasing doses of peanut allergen in order to decrease patients’ sensitivity to peanut allergen (ClinicalTrials.gov ID NCT01324401).^28^ Participants underwent a buildup phase where they were given incrementally increasing amounts of peanut protein, followed by a maintenance period with a daily dose of 4 grams. After 12 weeks of maintenance, participants underwent an oral food challenge (OFC1), followed by 1 month of peanut avoidance. A final double-blind, placebo controlled food challenge (OFC2) was performed to assess transient desensitization (those who failed OFC2) versus sustained unresponsiveness (those who passed OFC2). Blood sampling for metabolome datasets occurred at baseline, early build-up (at the peak of AEs), OFC1, and final OFC2 and was conducted for those with sufficient sample who developed transient desensitization or sustained unresponsiveness (n=20).

For all three cohorts, written parental consent on behalf of the child and child’s assent, if age appropriate, were obtained. All three studies were approved by the trial sites Institutional Review Boards. Further study design details are available in the original publications for each of the studies.^26–28^

### Food Allergy Case identification

*GACRS:* The parents of study participants were asked by questionnaire if their child experienced a food allergy and to which food, and we defined cases as parental report of an allergy to one of the 8 most common food allergens (milk, soy, egg, wheat, fish, shellfish, peanuts, tree nuts). Unaffected controls were those whose parents did not report food allergy. Participants with a report of a food allergy to an atypical allergen were excluded.

*VDAART:* Participants were given a detailed food allergy questionnaire which included questions about suspected food allergies, symptoms associated with reactions, positive testing by skin test or serum IgE test, and prescription with an epinephrine auto-injector. Some patients also had corroborating specific IgE levels for foods. Patients with a convincing history (report allergen triggering hives, swelling, cough, wheeze, vomiting, diarrhea) and evidence of sensitization (by positive blood IgE or reported positive skin or blood testing) were reviewed by an allergist and included as cases.

Unaffected controls were those who did not report food allergy, and anyone reporting a food allergy but not meeting criteria for diagnosis was excluded.

*PNOIT*: Inclusion criteria included objective symptoms after peanut ingestion, and either a peanut-specific IgE >10 kU/L or elevated peanut-specific skin prick test >8 mm. All patients demonstrated IgE-mediated symptoms during the early dosing of OIT, confirming the presence of true IgE-mediated food allergy.^28^

### Untargeted Metabolomics

Plasma samples were collected from all three populations, and metabolomic profiling was performed by the Clish laboratory at the Broad Institute for GACRS and PNOIT, and by Metabolon Inc for VDAART. Liquid chromatography coupled with tandem mass spectrometry (LC/MS/MS) methods were used to examine 4 classes of metabolites: 1) hydrophilic interaction liquid chromatography (HILIC)-MS/MS in positive ion mode (e.g., amino acids); (2) HILIC-MS/MS in negative ion mode (e.g., cAMP); (3) C8 reversed phase LC-MS/MS for lipids (e.g., steroids), and (4) C18 reversed phase LC-MS/MS for free fatty acids, (e.g., bile acids and eicosanoids). Features were indexed by their mass-to-charge ratio and LC retention time and metabolite identities were confirmed using known standards.^30, 31^ All features were log transformed to normalization and *pareto*-scaled to reduce the variation in fold change differences between the features. To ensure uniform classification of metabolite subclasses between the three cohorts, the Relational Database of Metabolic Pathways (RaMP) was used to identify the chemical subclass of each metabolite based on The Human Metabolome Database identifiers (HMDB v.5.0).^32, 33^

### Statistical Analysis

Comparisons of demographics were conducted using chi-square or Fisher’s exact tests, between individuals with food allergy and unaffected controls, and also between patients with sustained unresponsiveness versus transient desensitization. For GACRS and VDAART, we constructed generalized linear models for each cohort separately to assess the association of metabolites with reported food allergy, with adjustment for sex, age at blood collection, and for VDAART, race (Black, White, Asian, Other). For PNOIT, we used generalized linear regression (GLM) modeling, with adjustment for age, to identify significant metabolites, with linear regression to determine metabolites varying with time on OIT (from baseline to post-OIT), and logistic regression to identify metabolites associated with outcome (transient desensitization versus sustained unresponsiveness), adjusted for time on OIT from baseline to post-OIT. To summarize findings in PNOIT, we defined a set of key pathways of interest based on biological significance in atopic disease (polyunsaturated fatty acids, which include arachidonic acid metabolites, and sphingolipids), and based on top metabolite subclasses that were altered in the setting of food allergy compared to unaffected controls from VDAART & GACRS. The full set of findings are included in the corresponding supplemental figures for each set of analyses. Pathway analysis was performed using both chemical enrichment analysis (reported from the metabolite subclass) and biological pathway enrichment (based on biological pathways from KEGG from HMDB v5.0, Reactome v85, and WikiPathways v20230710) using the RaMP v2.3.0 (RaMP-DB R package).^32, 34–36^ All subclasses were defined using ClassyFire structural classification, from HMDB v5.0.^34, 37^ To capture the full range of potentially relevant subclasses and pathways, individual metabolites were reported based on a nominal significance of p<0.05, while chemical subclasses and biological pathways were reported using FDR corrected significance of p<0.05. All analyses were performed using R 4.2.2.^38^ Metabolomic data are available through the dbGaP for GACRS, the ECHO consortium for VDAART, and Metabolomics Workbench for PNOIT.^39–41^

## RESULTS

### Population demographics of three studies: two observational cohorts, GACRS and VDAART, and the PNOIT interventional trial

We used 2 observational cohorts, GACRS (n=1040 children with asthma) and VDAART (n=384 healthy infants) to study the metabolomics of food allergy. The prevalence of reported food allergy in GACRS was 5% (57/1040), while the prevalence of reported, validated, food allergy in VDAART was 6% (22/384) (**Table 1**). Both populations were approximately 40% female; differences in racial/ethnicity demographics were consistent with those between the countries of each study. Regarding atopic disease, the GACRS cohort had higher proportions of allergic rhinitis, whereas, the VDAART cohort had higher percentages of eczema, consistent with the older versus younger ages of each cohort. Approximately a quarter of the VDAART cohort experienced asthma/persistent wheeze by age 3, compared with all reporting asthma symptoms in GACRS due to study design.

**Table 1:** Demographics of GACRS, VDAART, and PNOIT studies. For GACRS, race was not collected from the Costa Rican population.

| **Genetics of Asthma in Costa Rica Study (GACRS)** |  |  |  |  |
| --- | --- | --- | --- | --- |
|  | Overall<br>N=1040 | Food Allergic<br>(n=57) | Unaffected<br>Controls (n=983) | p-value |
| **Male (%)** | 619 (60%) | 28 (49%) | 591 (60%) | 0.13 |
| **Age at Sampling**<br>**(years)** | 9.2 (1.9) | 9.0 (1.8) | 9.3 (1.9) | 0.39 |
| **Hispanic/Latinx** | 1040 (100%) | 57 (100%) | 983 (100%) | 1.00 |
| **Physician**<br>**Diagnosed** |  |  |  |  |
| **Asthma** | 1012 (97%) | 55 (97%) | 957 (97%) | 1.00 |
| **Allergic Rhinitis** | 335 (32%) | 24 (42%) | 311 (32%) | 0.14 |
| **Eczema** | 45 (4%) | 6 (11%) | 39 (4%) | 0.04 |
| **Vitamin D Antenatal Asthma Reduction Trial (VDAART)** |  |  |  |  |
|  | Overall<br>N=384 | Food Allergic<br>(n=22) | Unaffected<br>Controls (n=362) | p-value |
| **Male (%)** | 207 (54%) | 12 (55%) | 195 (54%) | 1.00 |
| **Age at Sampling**<br>**(years)** | 3.0 (0.09) | 3.0 (0.1) | 3.0 (0.09) | 0.30 |
| **Race** |  |  |  | 0.11 |
| **Black** | 186 (48%) | 16 (73%) | 170 (47%) |  |
| **White** | 125 (33%) | 4 (18%) | 121 (33%) |  |
| **Asian** | 22 (6%) | 0 (0%) | 22 (6%) |  |
| **Other** | 51 (13%) | 2 (9%) | 49 (14%) |  |
| **Hispanic/Latinx** | 126 (33%) | 7 (32%) | 119 (33%) | 1.00 |
| **Reported Asthma/**<br>**Wheeze by age 3** | 100 (26%) | 15 (68%) | 85 (24%) | <0.001 |
| **Physician Diagnosed** |  |  |  |  |
| **Asthma** | 53 (14%) | 12 (55%) | 41 (11%) | <0.001 |
| **Allergic Rhinitis** | 72 (19%) | 11 (50%) | 61 (17%) | <0.001 |
| **Eczema** | 184 (48%) | 22 (100%) | 162 (45%) | <0.001 |
| **Peanut Oral Immunotherapy Trial (PNOIT)** |  |  |  |  |
|  | Overall<br>N=20 | Sustained<br>Unresponsiveness<br>(SU) (n=9) | Transient<br>Desensitization<br>(TD) (n=11) | p-value |
| **Male (%)** | 15 (75%) | 7 (78%) | 8 (73%) | 1.00 |
| **Mean Age at**<br>**Enrollment (years**<br>**(SD))** | 9.9 (2.1) | 9.8 (2.3) | 10.0 (2.1) | 0.81 |
| **Race** |  |  |  | 0.40 |
| **Asian** | 2 (10%) | 1 (11%) | 1 (9%) |  |
| **White** | 16 (80%) | 8 (89%) | 8 (73%) |  |
| **Other** | 2 (10%) | 0 (0%) | 2 (18%) |  |
| **Hispanic/Latinx** | 1 (5%) | 0 (0%) | 1 (9%) | 1.00 |
| **Allergic comorbidities** |  |  |  |  |
| **Asthma** | 8 (40%) | 3 (33%) | 5 (45%) | 0.93 |
| **Allergic Rhinitis** | 12 (60%) | 7 (78%) | 5 (45%) | 0.31 |
| **Eczema** | 11 (55%) | 3 (33%) | 8 (73%) | 0.19 |

From the original PNOIT trial cohort,^28^ 30 participants were enrolled, 27 completed treatment, and 13 participants developed transient desensitization (TD) compared to 9 with sustained unresponsiveness (SU), and 5 participants experienced treatment failure. Metabolomic profiling was performed on 11 of 13 participants found to have transient desensitization (TD) and all 9 participants with sustained unresponsiveness (SU); 2 samples from participants who developed TD were not included in metabolomic profiling due to inadequate sample volumes. All participants in the PNOIT interventional trial had IgE-mediated food allergy by study design, and rates of asthma and allergic rhinitis were similar to those seen in the food-allergic population of VDAART (**Table 1**).

### Food allergy in GACRS and VDAART associated with alterations in bile acid and histidine metabolism

After quality control measures, 595 named metabolites in GACRS and 655 in VDAART were available for analysis. Regression analysis revealed that 16 (3%) metabolites in GACRS and 17 (3%) metabolites in VDAART were associated with food allergy (**Figure 2**). Among these metabolites, the most common subclasses included amino acids (11 metabolites) and bile acids (8 metabolites). Within the amino acid pathway, histidine metabolism was the most prominent, with 2 metabolites increased among individuals with food allergy individuals (N-acetyl histidine and urocanic acid, (range of odds ratios (OR) 1.70-4.08, and range of p-values 0.017-0.018), and one metabolite, ergothioneine (OR 0.17, p=0.023) increased in unaffected controls. Among bile acids, metabolites included both primary and secondary bile acids, and all 8 were increased in those with food allergy (range of OR: 1.13-3.44, range of p-values: 0.0002-0.04). Based on these findings, we included bile acids and histidine metabolites in our list of curated pathways to study in OIT.

**Figure 2:**
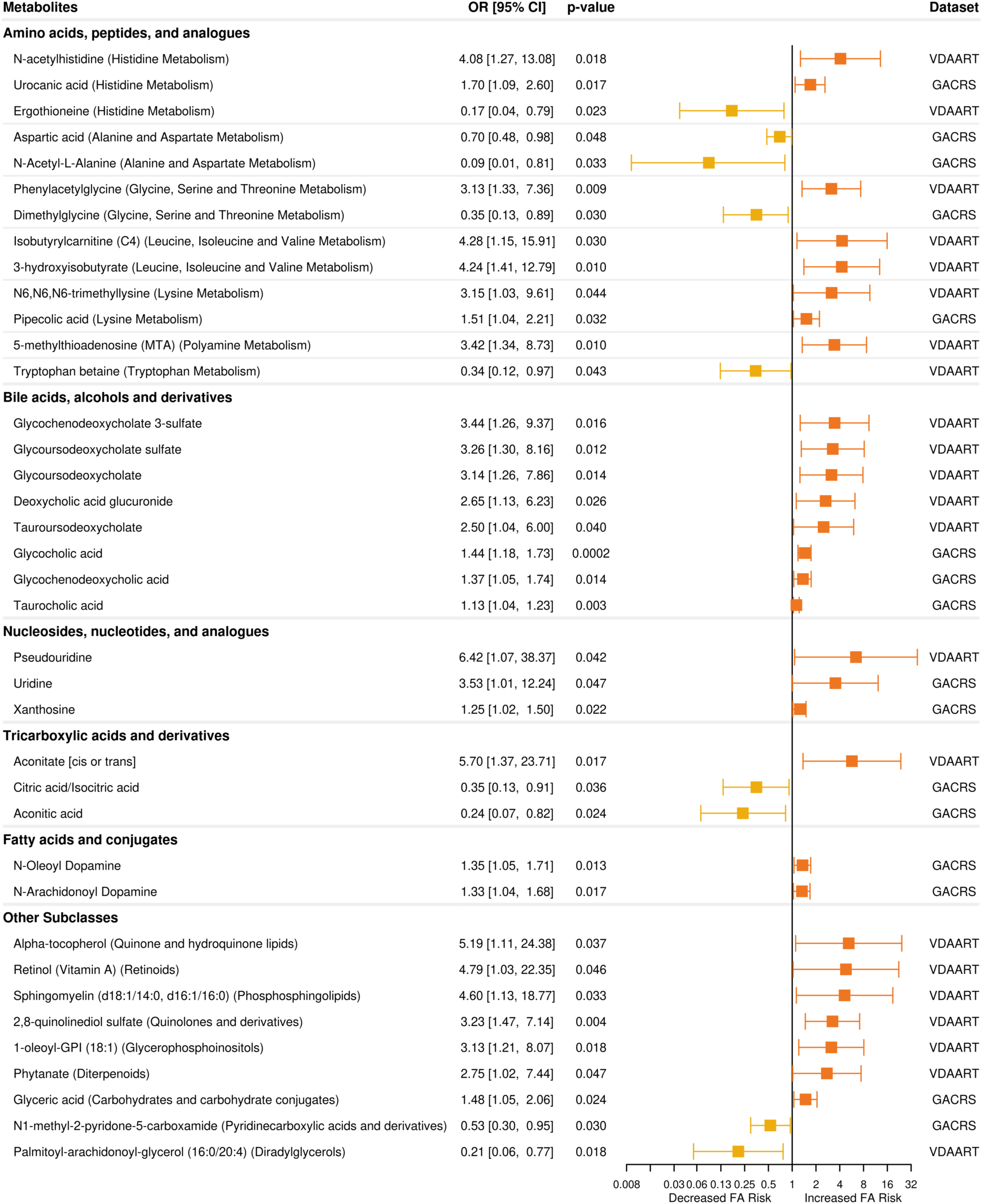
Metabolomic profiling of Food Allergy in the GACRS and VDAART pediatric populations identifies altered bile acid and histidine metabolic pathways. Plasma metabolites associated with food allergy using logistic regression within each cohort, adjusted for age, sex, and race (for VDAART only), identified 29 significant metabolites higher in participants with food allergy (orange) and 9 significant metabolites higher in unaffected controls (yellow). Pathways with 3 or more unique metabolites included bile acids and histidine metabolism. Metabolomic profiling for each cohort was conducted in different laboratories, leading to some differences in annotation of metabolites.

### Polyunsaturated fatty Acids (PUFAs), including arachidonic acid derivatives, decrease over time on OIT in all participants (PNOIT)

After quality control measures, 529 named metabolites were available for analysis in the PNOIT cohort. Analyzing metabolite trajectories over time, metabolites showed longitudinal differences associated with OIT among all participants (**Figure E1**). Subclasses with the highest number of significant metabolites included eicosanoids (17 significant metabolites decreased over time), glycerophosphocholines (14 significant metabolites increased and 2 decreased over time), and glycerophosphoethanolamines (9 significant metabolites increased and 1 decreased over time) (**Figure 3a**). Several subclasses were notable in that all significant metabolites (and a majority of non-significant metabolites) in that class changed in the same direction, including increasing triacylglycerols, decreasing linoleic acids, and decreasing eicosanoids over time on OIT. Chemical subclass pathway enrichment of the significant metabolites identified top subclasses including eicosanoids, (q=2.4×10^−20^, enrichment ratio=43), glycerophosphocholines (q=1.4×10^−13^, enrichment ratio=15), and steroid esters (q=3.7×10^−6^, enrichment ratio=33) (**Figure 3b**). Biological pathway enrichment analysis also identified a number of PUFA/eicosanoid related pathways, including arachidonic acid oxylipin metabolism (q=7.6×10^−5^, 13% of metabolites in pathway), metabolism of alpha-linolenic acid (q=1.1×10^−5^, 35% of pathway), eicosanoid metabolism via lipooxygenases (LOX) (q=0.00072, 19% of pathway), and eicosanoid synthesis (q=0.02, 15% of metabolites in pathway). Additionally, amino acids: histidine catabolism (q=4.0×10^−5^, 35% of pathway), and sphingolipid metabolism (q=0.025, 21% of pathway) were also identified by biological pathway enrichment.

**Figure 3.**
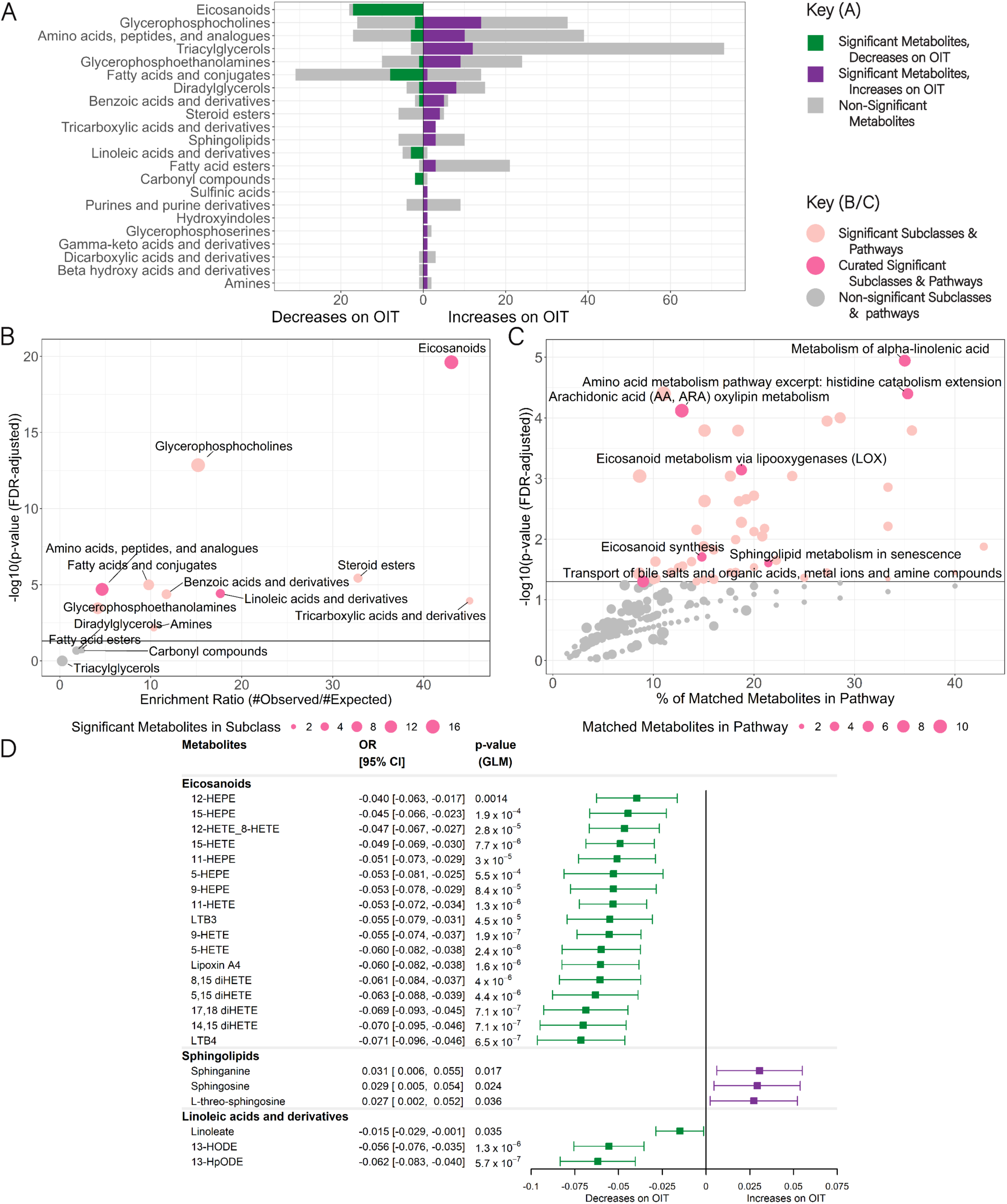
– PNOIT: Metabolites associated with change over time in the entire cohort during OIT. (A) Subclasses of significant (p<0.05) metabolites with the number of metabolites increasing (purple) and decreasing (green). The total number of detected non-significant metabolites within each subclass is denoted in gray. Pathway enrichment analyses show (B) enrichment ratios (ratio of observed significant metabolites divided by expected significant metabolites) of chemical subclasses plotted by the FDR adjusted p-value plotted on a log10 scale, and (C) percent of significant metabolites divided by the total metabolites within the pathway plotted by the FDR-adjusted p-value plotted on a log10 scale. In B & C, black line denotes q<0.05, and key pathways of interest are labeled in magenta (bile acids, eicosanoids, linoleic acids, sphingolipids, histidine metabolites). (D) Association of each metabolite with time on OIT.

Based on biological significance in other atopic diseases and key metabolites associated with food allergy in GACRS and VDAART, we defined a set of key pathways of interest, including histidine metabolism, bile acids, sphingolipids, and polyunsaturated fatty acids (PUFAs) including linoleic acid derivatives and eicosanoids (**Figure 3d**, all significant metabolites in **Figure E1b**). Of note, bile acids were not detected in association with asthma in the VDAART cohort. Among PUFAs, significant decreases in all participants during OIT were detected among the eicosanoids, including leukotriene B3 and B4, lipoxin A4, and several HETEs/DiHETES (range of coefficients: –0.04 to –0.071, range of p values:1.9×10^−7^ to 0.0014). All linoleic acid derivatives, including linoleic acid, 13-HpODE, and 13-HODE decreased (range of coefficients: –0.062 to –0.015, range of p-values: 5.7×10^−07^ to 0.035) and all detected sphingolipids increased over time on OIT (range of coefficients: 0.027 to 0.031, range of p values: 0.017 to 0.036). No bile acid or main histidine pathway metabolites were identified as changing over time on OIT.

### Enrichment of bile acid, histidine, and eicosanoid pathways in sustained unresponsiveness in PNOIT

Adjusting for time, 116 named metabolites (21.9% of total named metabolites) differed between individuals on OIT with sustained unresponsiveness compared to transient desensitization (**Figure E2a**). Top subclasses, based on number of metabolites observed, included glycerophosphocholines (8 higher in SU and 10 higher in TD, q=6.8×10^−16^; enrichment ratio=18) and amino acids, peptides, and analogues (12 higher in SU and 2 higher in TD, q=7.9×10^−7^; enrichment ratio=6) (**Figure 4a**). Subclasses that were notable because all significant metabolites (and a majority of non-significant metabolites) in that class were higher in one group included all triacylglycerols higher in transient desensitization (6 metabolites), and all eicosanoid metabolites (8 metabolites) higher in sustained unresponsiveness. Chemical enrichment analysis identified differences in bile acid (q=4.1×10^−8^; enrichment ratio=33) and eicosanoid (q=7.9×10^−7^; enrichment ratio=19) subclasses between the two groups (**Figure 4b**), and biological pathway enrichment (**Figure 4c**) showed significant enrichment of several bile acid pathways, including recycling of bile acids and salts (q=2.0×10^−7^; 45% of pathway), transport of bile salts (q=5.4×10^−5^; 15% of pathway), synthesis of bile acids and bile salts (q= 0.00019; 18% of pathway), and primary bile acid synthesis (q=0.017; 17% of pathway), as well as, eicosanoid metabolism via lipooxygenases (LOX) (q=0.030; 9% of pathway), histidine catabolism (q=0.015; 14% of pathway), and sphingolipid metabolism (q=0.004; 9% of pathway). Among the curated set of pathways of interest, histidines (1 higher in SU, 1 with TD), bile acids (2 higher in SU and 5 with TD), eicosanoids (8 higher in SU), and linoleic acid metabolites (1 higher in TD) all contained metabolites significantly associated with clinical outcome (**Figure 4d** & all significant metabolites in **Figure E2b**).

**Figure 4.**
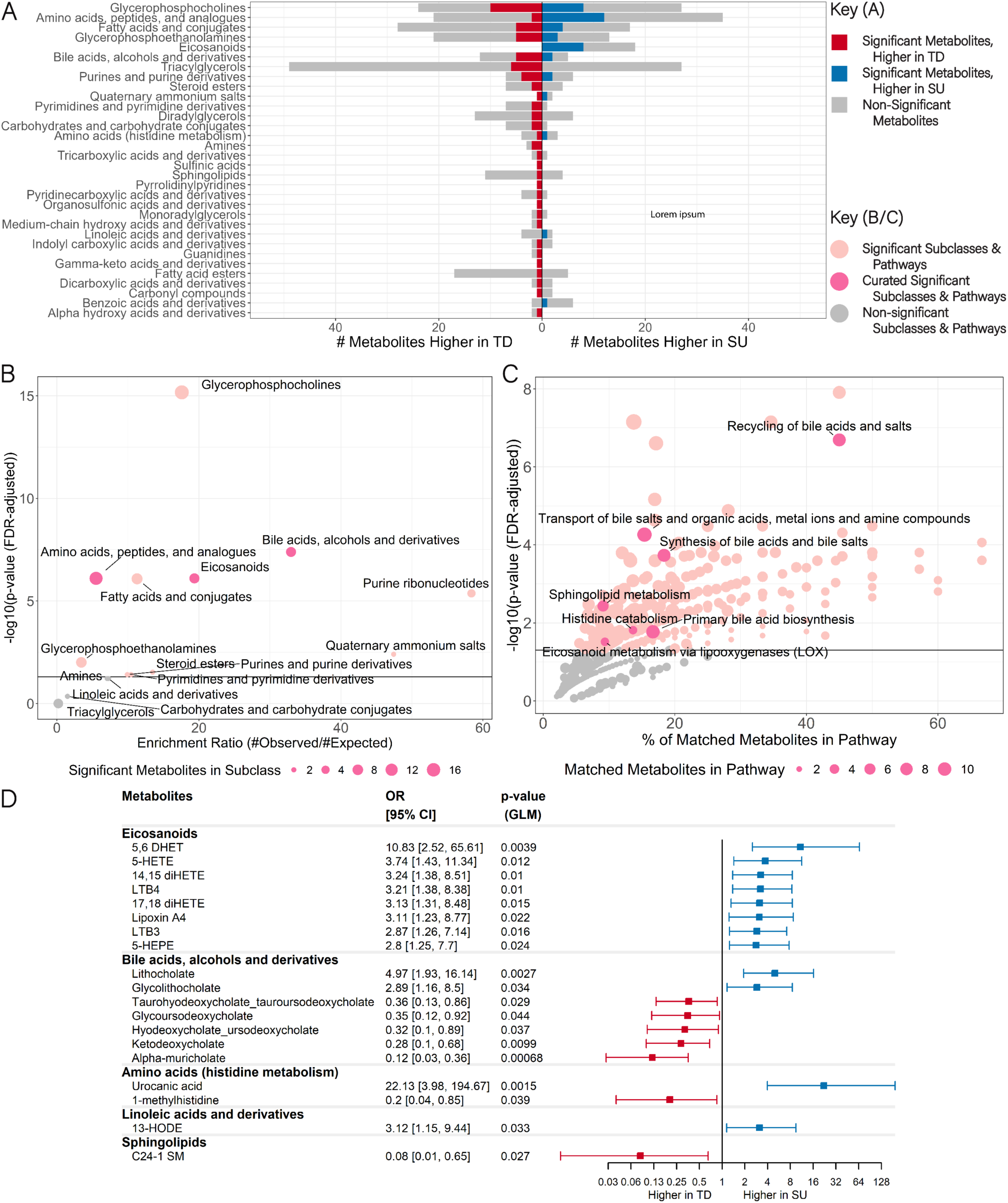
– PNOIT: Metabolites differing between participants who develop sustained unresponsiveness (SU) compared to transient desensitization (TD). (A) Subclasses of significant (p<0.05) metabolites with the number of metabolites higher in either SU (blue) or TD (red). The total number of detected non-significant metabolites within each subclass is denoted in gray. Pathway enrichment analyses show (B) the ratio of observed significant metabolites divided by the number of expected metabolites within the chemical subclass plotted by the FDR-adjusted p-value plotted on a log10 scale, and (C) the percent of significant metabolites divided by the total metabolites within the biological pathway plotted by the FDR-adjusted p-value plotted on a log10 scale. In B & C, black line denotes q<0.05, and key pathways of interest are labeled in magenta (bile acids, eicosanoids, linoleic acids, sphingolipids, histidine metabolites). (D) Association of each metabolite with time on OIT.

Among bile acids, most were higher in transient desensitization (range of OR: 0.12-0.36, range of p=0.00068-0.044), with two in particular higher in sustained unresponsiveness, lithocholate (OR: 4.97, p = 0.0027) and glycolithocholate (OR: 2.89, p = 0.034) (**Figure 4b**). For some metabolites, ursodeoxycholate, alpha-muricholate, and ketodeoxycholate, trajectories differed at baseline, with higher levels among transiently desensitized individuals, and converged over time on OIT (**Figure 5a**). For lithocholate, levels stayed relatively stable during OIT, with higher concentrations among participants with sustained unresponsiveness, whereas the downstream metabolite glycolithocholate demonstrated converging trajectories during OIT.

**Figure 5.**
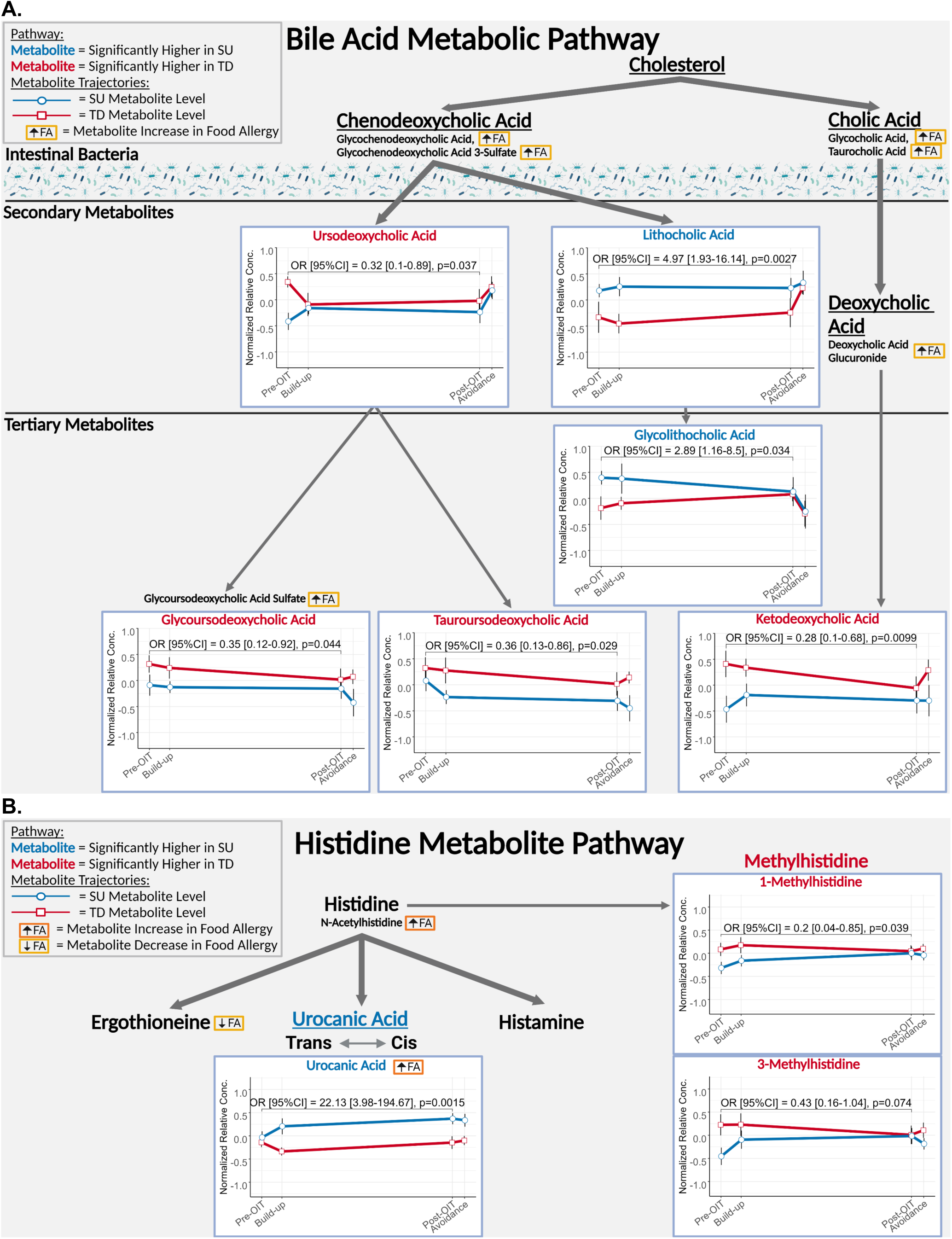
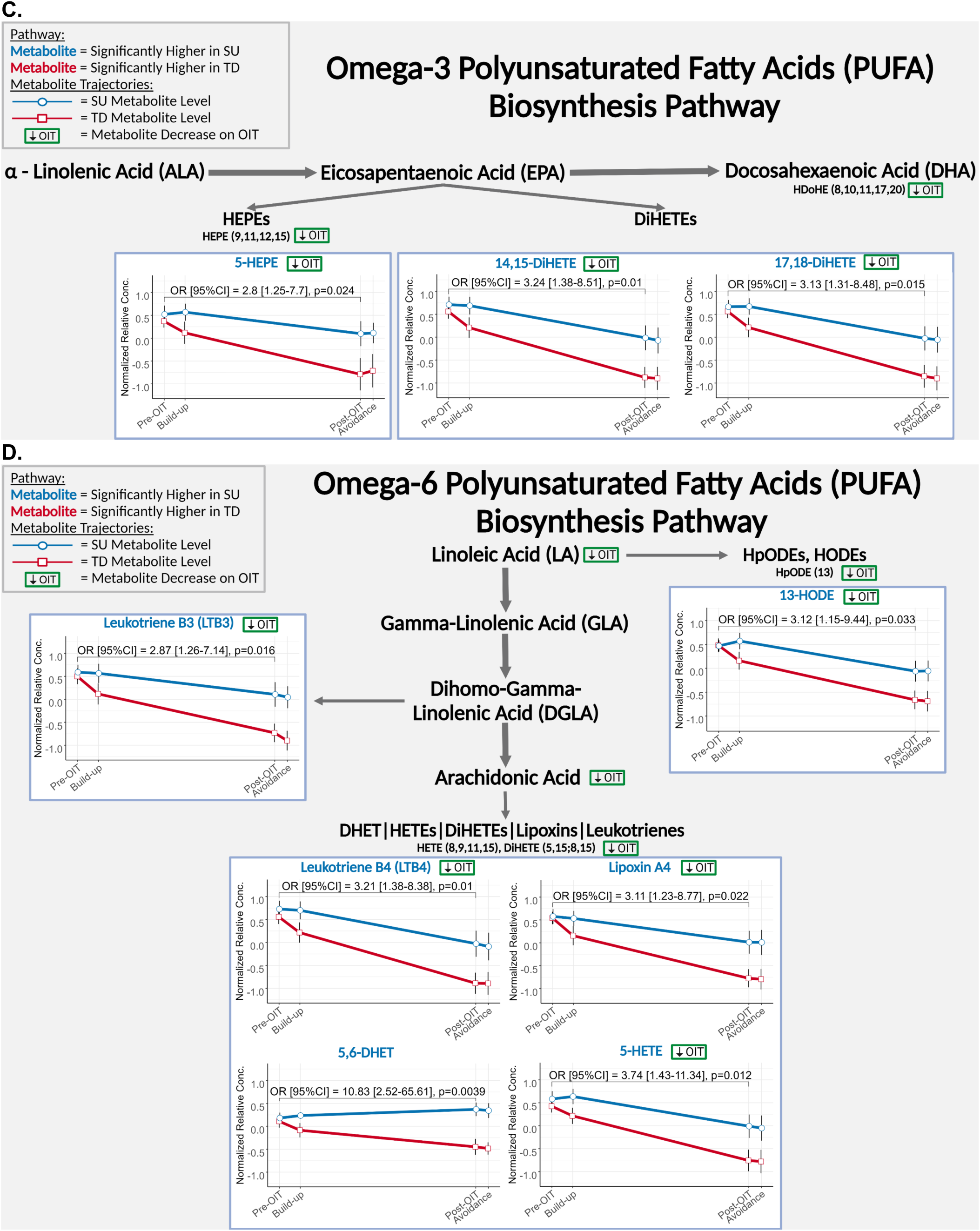
– PNOIT: Overview of key metabolic pathways of interest and metabolite trajectories during OIT, between transient desensitization (TD, red) and sustained unresponsiveness (SU, blue). A. Bile acid pathway. B. Histidine Pathway. C. Omega 3 fatty acids (PUFAs). D. Omega 6 fatty acids (PUFAs). Metabolites associated with food allergy are denoted in small boxes colored by orange (higher in food allergy) or yellow (higher in unaffected controls). Metabolites associated with time on OIT in the entire cohort are denoted in small boxes in green (decreasing during OIT) or purple (increasing during OIT). Modified derivatives of the main metabolite are listed under the main metabolite. Only metabolites that significantly differ in association with food allergy and/or OIT are depicted, with major biosynthetic processes in thick arrows and simpler metabolic modifications in thin arrows. Graphs are only displayed for metabolites that differed significantly between SU & TD. Listed odds ratio and confidence interval (OR [95%CI]) statistics are derived from logistic regression models analyzing the association of the metabolite with OIT outcome (SU versus TD), adjusted for age and time on OIT.

Among PUFAs, significant metabolites were detected from the eicosanoid subclass, including leukotriene B_3_ (LTB_3_) and LTB_4_, lipoxin A_4_ (LXA_4_), 5,6-DHET (5,6-DiHETrE), and several HETEs/DiHETES (range of OR: 2.80-10.83, range of p: 0.0039-0.024) (**Figure 4b**). With the exception of 5,6-DiHETrE/5,6-DHET, all eicosanoid metabolites decreased over time on OIT, with more dramatic decreases among those participants with transient desensitization than with sustained unresponsiveness (**Figure 5b**).

For histidine metabolism, the 1-methylhistidine was higher in subjects with transient desensitization (OR: 0.20, p=0.039), whereas urocanic acid (OR: 22.13, p=0.0015), was higher among participants with sustained unresponsiveness (**Figure 4b**). The histidine derivatives demonstrated higher concentrations at baseline and converged over time on therapy (**Figure 5c**). Meanwhile, urocanic acid levels were similar between both groups of participants at baseline, and then diverged during OIT, most dramatically between baseline and buildup, with higher levels among participants with sustained unresponsiveness.

## DISCUSSION

In this analysis of data from three studies including an international cohort, we conducted untargeted metabolomic profiling of two epidemiologic cohorts and one interventional clinical trial to identify metabolites associated with IgE-mediated food allergy and clinical responses to PNOIT. To our knowledge, this is the first study of metabolomics in the setting of food allergen immunotherapy in humans. In this analysis, we discovered a number of key metabolites (including bile acids and histidine metabolites) associated with parental report of food allergy in GACRS, as well as with IgE-mediated food allergy in VDAART. In PNOIT, exploration of these pathways among participants with confirmed IgE-mediated food allergy demonstrated that bile acids, eicosanoids, and histidine metabolites were associated with differential responses to OIT, distinguishing individuals with transient desensitization to OIT lasting only the duration of active treatment from those with sustained unresponsiveness with protection lasting at least a month beyond the cessation of OIT.

## The immunomodulatory roles of bile acids in food allergy and OIT

### Elevated bile acids in food allergy

Our work showed significantly higher levels of bile acids among children with food allergy compared to unaffected controls, replicating findings from another study of serum metabolomics in food allergy.^14^ These bile acids however, were not associated with other atopic diseases, such as asthma in the VDAART cohort. Other work in VDAART has shown that stool from children with food allergy is notable for lower levels of bile acids compared to both unaffected controls and sensitized children (those with food specific antibodies but not food-allergy).^13, 18^ This work has been replicated in another study showing not only lower bile acids in stool from children with food allergy compared to healthy controls, but also lower levels of bile acids in children with persistent food allergy.^42^ Combined with our findings that children with food allergy in VDAART had higher levels of bile acids in the plasma, this suggests the increase in plasma bile acids is most likely due to increased bile acid reabsorption in food-allergy, rather than increased overall metabolism of bile acids by gut microbiota. Increased bile acid absorption has been implicated in the mechanism of liver diseases and type 2 diabetes mellitus, and increased bile acid levels have been linked to increases in intestinal permeability, notable because IgE-mediated food allergy is also marked by dysfunctions in intestinal permeability.^43, 44^

### Lower bile acids & and higher lithocholate derivatives in sustained efficacy on OIT

In addition to elevated levels in food allergy, our work shows generally higher levels of bile acids, especially deoxycholate derivatives, among individuals who only develop transient desensitization on OIT (potentially a more severe phenotype of food allergy) (**Figure 5a**). The exceptions to this pattern, lithocholate & glycolithocholate are higher among children who later develop sustained unresponsiveness after stopping therapy. Given lithocholate derivatives actively alter T cell differentiation,^45, 46^ our data suggest that lithocholate is not only a biomarker but may actively increase the likelihood of allergic remission. These bile acid profiles seem to be present even before the initiation of OIT, and while some bile acid trajectories converge over time on OIT, others, like lithocholate remain relatively stable.

### Immunomodulatory roles of lithocholate on the T cell subsets associated with OIT outcomes

Recent findings have uncovered many novel roles for bile acids in immune regulation of T cells.^47^ Bile acids are synthesized from cholesterol in the liver, secreted into the intestine to help with digestion, modified by gut bacteria into secondary bile acids and then reabsorbed.^48^ In murine models, a screen of primary and secondary bile acids discovered that two lithocholate metabolites blocked Th17 differentiation and function and enhanced Treg differentiation, stability, and function.^45, 46^ Colonization of mice with human bacterial strains that produce certain lithocholate derivatives can suppress Th17 levels and increase the gut Treg population. ^46, 49^ In human studies of OIT, decreases in peanut-specific Th17 frequency have been observed between those who achieve desensitization compared to those who fail treatment,^15^ as well as decreased overall Th17 frequency over time on OIT among older children and teens.^50^ While peripheral allergen-specific Treg frequencies do not differ by sustained unresponsiveness outcomes in OIT,^15, 51^ mucosal allergen-specific Treg responses have not been studied, and OIT has been shown to induce increased expansion of the overall Treg compartment in patients who develop sustained unresponsiveness, compared to those who develop transient desensitization.^52, 53^ We propose that bile acids, such as lithocholate derivatives, modulate T cell percentages and function to increase the likelihood of remission in OIT. Overall, these findings may reflect not only the microbial differences in individuals who develop longer lasting protection from OIT, but also an active role for bile acids in mediating these immunopathogenic differences.

## Altered histidine metabolism in food allergy and OIT

Our work demonstrated altered serum histidine metabolism in food allergy, with metabolites such as urocanic acid (UCA) and histidine were increased in those with food allergy while ergothioneine was increased among unaffected controls. Others have also found altered histidine metabolism associated with food allergy and food allergy development in both the serum and stool.^14, 20^ Histidine is an essential amino acid that can be converted into both inflammatory mediators (e.g. histamine) but also anti-inflammatory derivatives such as the cis-form of urocanic acid (UCA). In the skin, peanut allergic children had lower levels of cis-UCA compared to non-atopic controls, independent of their atopic dermatitis.^54^ Given these findings, the elevation of UCA in food allergic children in GACRS is unexpected, but may reflect differences between the isomers or between the plasma and skin metabolome.

These data also demonstrate a role for histidine metabolites in OIT responses. In particular urocanic acid (UCA) levels are higher in individuals with sustained unresponsiveness in PNOIT, though that difference was apparently induced by OIT (**Figure 5b**). UCA has a number of immunomodulatory functions, including increasing IL-10 expression, decreasing IFN-gamma production by antigen specific T cells, and increasing Treg percentages.^55, 56^ IL-10 producing cells (including T regs, CD4+ T cells, and regulatory B cells) are important for the development of allergic tolerance.^8^ Furthermore, our data revealed higher levels of succinic acid in our participants with transient desensitization (**Figure E2a, Dicarboxylic Acids & Derivatives**), and succinic acid inhibits histidase, the enzyme that converts histidine into urocanic acid.^57^ Both succinic acid and urocanic acid creation and degradation are affected by skin and gut microbial flora,^57, 58^ all of which supports another potential pathway by which the microbiome interacts with the immune system to effect tolerance on immunotherapy. Given the known bidirectional nature of microbial-immune cross-talk,^59, 60^ this may provide a potential explanation for why changes in urocanic acid do not occur until after OIT is initiated. Additionally, UCA is a breakdown product of filaggrin, a key epithelial barrier protein which is a risk factor for food allergy and is decreased in atopic dermatitis and eosinophilic esophagitis (EoE) but normalizes after treatment of EoE.^61–63^ Given the links between EoE, tissue eosinophilia, and OIT, the increase in UCA in sustained unresponsiveness may reflect changes in filaggrin expression during OIT.^64^ Overall, the divergence in UCA trajectories between sustained unresponsiveness and transient desensitization suggest that the role of UCA, as an immunomodulatory metabolite or as a reflection of changes in the microbiome or filaggrin, merits further study.

## Decreases in PUFAs during OIT

PUFAs (often described as omega-3 and omega-6 fatty acids) encompass a variety of subclasses of metabolites, including eicosanoids and linoleic acid derivatives. Of these groups, eicosanoids (which include arachidonic acid derivatives) have a well described role in the pathophysiology of allergic disease, with many having an allergic pro-inflammatory role, including leukotrienes, and prostaglandins.^65^ In our data, all measured eicosanoids and linoleic acid derivatives decreased over time on OIT, and we hypothesize that the general decrease in these inflammatory mediators is a result of reduced inflammatory circuits and successful treatment (**Figure 5c**). The finding that participants with transient desensitization exhibited more dramatic decreases than those with sustained unresponsiveness was unexpected. One potential explanation involves a key effector CD4 T cell subset, referred to as pathogenic effector (peTh2)^66^ or Th2A,^67^ which has emerged as a likely regulator of multiple allergic disease, including food allergy. These Th2A are notable for several features that distinguish them from Th2-skewed T follicular helper cells (necessary for high-affinity IgE) and other Th2 subsets, including production of IL-9, expression of the transcriptional regulator PPARγ, and the marked upregulation of a set of eicosanoid sensing and synthesizing genes. ^68–72^ We and others have shown that Th2A are suppressed by OIT in both TD and SU.^15, 51^ Recent data from cellular analysis of young toddlers undergoing OIT also demonstrated that Th2A cells were suppressed over time by OIT, and suggests one reason a decrease may be more dramatic in TD versus SU. ^51^ For the one subject analyzed who achieved SU, there were few Th2A cells present at baseline.^51^ Another possibility is that these trajectories represent a shift between the arms of the eicosanoid pathway, where patients who go on to develop sustained unresponsiveness, have a more dramatic decrease in prostaglandin production. Prostaglandins were not detected in the plasma from these samples, but this merits further study in future trials, ideally by the analysis of urine metabolites.

## Limitations

This analysis compares three remarkably disparate populations: a cohort from Costa Rica, a country with high prevalence of allergic asthma, a healthy cohort of US infants born to pregnant mothers from an interventional trial of Vitamin D supplementation during pregnancy, and finally, children with food allergy treated with peanut OIT. These cohorts include ages ranging from toddlers to teenagers, and children with varied allergic comorbidities. The diagnosis of IgE-mediated food allergy in these cohorts was of variable stringency, with the PNOIT cohort having the most reliable diagnosis (confirmed by clinician observed reactions to allergen), the VDAART population being most similar to a standard clinical cohort of IgE-mediated food allergy (diagnosed based on a combination of sensitization and history, but often missing confirmatory food challenges) and GACRS cohort being the loosest, relying on parentally reported food allergy. This work also faces limitations in the small sample sizes of participants with food allergy. Because samples were being processed in parallel for isolation of cells, samples were not handled with strict temperature control required for ideal metabolomics processing prior to freezing. This may have altered detection of certain metabolites. Furthermore, the metabolomics profiling was conducted by two different groups, which is known to influence the uniformity of metabolite detection, particularly notable in the replication of pathways between VDAART and GACRS but not generally the exact metabolites. Despite these differences in study population and metabolomics processing, we found replication of pathways between cohorts. Also our work supports findings in the food allergy literature showing differential production of sphingolipids, tryptophan metabolites, alanine metabolites, glycerophosphoinositols, and pyridine carboxylic acid derivatives (also described as Vitamin B3 metabolites).^14, 73^ Furthermore, many of the pathways associated with food allergy were differentially produced when comparing the two clinical responses to OIT, adding robustness to the hypothesis that these metabolites represent an important part of the immune pathogenesis of IgE-mediated food allergy.

## Conclusion

In summary, we have identified key metabolite pathways associated with food allergy and differential responses to OIT. Many of these metabolites, including bile acids, eicosanoids, and histidine/urocanic acid, have active roles in modulating the same T cell subsets that are thought to mediate remission in allergen immunotherapy. Further functional validation of these metabolic pathways in the context of allergic tolerance may both help identify patients most likely to benefit from OIT and guide the development of adjunctive therapies that can improve outcomes of allergen immunotherapy.

## Clinical Trial or Protocol number associated with this study

NCT01324401 (PNOIT), NCT00920621 (VDAART)

## Funding/Acknowledgments

The Genetics of Asthma in Costa Rica Study was supported by the National Institutes of Health (Grants HL066289, P01HL13225, R01HL123915). Vitamin D Antenatal Asthma Reduction Trial (VDAART) was supported by R01HL123546, R01HL091528, U01HL091528, and UH3ODO023268 from the National Heart, Lung, and Blood Institute (NHLBI); U54TR001012 from the National Centers for Advancing Translational Sciences; and U19AI168643 from the NIAID. GACRS & VDAART metabolomic profiling and additional analyses were supported by R01HL123915, R01HL141826, and R01HL155742. The Peanut Oral Immunotherapy (PNOIT) study clinical trial work was performed at the Harvard Clinical and Translational Science Center supported by Grant Numbers 1UL1TR001102, 8UL1TR000170 from the National Center for Advancing Translational Science, and 1UL1 RR025758 from the National Center for Research Resources. PNOIT metabolomics profiling was supported by the 2014 ARTrust Minigrant. This work is also supported by NIH NIAID (grant K23-AI130408 for YVV, T32AI007062 for JNS, 1R01AI155630 for SUP), and NHLBI (K01 HL146980 for RSK). CEW was supported by the Swedish Heart-Lung Foundation (HLF 20230463, HLF 20210519). SB is supported by the NIH NIAID (UM1 AI173380, U19 AI136053, R01 AI147028, U01 AI160082). EB is supported by NIEHS (P42ES027704) and NIGMS (R01GM141277 and RM1GM145416).

## Data Availability

Data are available through the dbGaP for GACRS study and the ECHO consortium for VDAART study. Data for the PNOIT study will be made available upon publication at Metabolomics Workbench.

