## Supplemental Figures and Tables for "Metabolomics of IgE-Mediated Food Allergy and Oral Immunotherapy Outcomes based on Metabolomic Profiling"

**Figure E1 – PNOIT: Metabolites associated with time on OIT**. Generalized linear model of metabolites significantly changing over time on OIT (increasing in purple and decreasing in green) adjusted for age, with (A) all significant metabolites, and (B) all identified metabolites in the key pathways of interest (bile acids, histidine metabolites, sphingolipids, eicosanoid & linoleic acid metabolites).

| Metabolites | OR [95% CI] | p-value (GLM) |
|---|---|---|
| **Eicosanoids** | | |
| 12-HEPE | -0.04 [-0.063, -0.017] | 0.0014 |
| 15-HEPE | -0.045 [-0.066, -0.023] | $1.9 \times 10^{-4}$ |
| 12-HETE_8-HETE | -0.047 [-0.067, -0.027] | $2.8 \times 10^{-5}$ |
| 15-HETE | -0.049 [-0.069, -0.03] | $7.7 \times 10^{-6}$ |
| 11-HEPE | -0.051 [-0.073, -0.029] | $3 \times 10^{-5}$ |
| 5-HEPE | -0.053 [-0.081, -0.025] | $5.5 \times 10^{-4}$ |
| 9-HEPE | -0.053 [-0.078, -0.029] | $8.4 \times 10^{-5}$ |
| 11-HETE | -0.053 [-0.072, -0.034] | $1.3 \times 10^{-6}$ |
| LTB3 | -0.055 [-0.079, -0.031] | $4.5 \times 10^{-5}$ |
| 9-HETE | -0.055 [-0.074, -0.037] | $1.9 \times 10^{-7}$ |
| 5-HETE | -0.06 [-0.082, -0.038] | $2.4 \times 10^{-6}$ |
| Lipoxin A4 | -0.06 [-0.082, -0.038] | $1.6 \times 10^{-6}$ |
| 8,15 diHETE | -0.061 [-0.084, -0.037] | $4 \times 10^{-6}$ |
| 5,15 diHETE | -0.063 [-0.088, -0.039] | $4.4 \times 10^{-6}$ |
| 17,18 diHETE | -0.069 [-0.093, -0.045] | $7.1 \times 10^{-7}$ |
| 14,15 diHETE | -0.07 [-0.095, -0.046] | $7.1 \times 10^{-7}$ |
| LTB4 | -0.071 [-0.096, -0.046] | $6.5 \times 10^{-7}$ |
| **Glycerophosphocholines** | | |
| C40-10 PC | 0.039 [0.019, 0.059] | $2.9 \times 10^{-4}$ |
| C36-5 PC plasmalogen-A | 0.032 [0.013, 0.052] | 0.0021 |
| C22-5 CE | 0.031 [0.012, 0.049] | 0.0017 |
| C38-6 PC | 0.026 [0.011, 0.042] | 0.0016 |
| C40-9 PC | 0.026 [0.011, 0.041] | 0.0013 |
| C38-7 PC plasmalogen | 0.026 [0.01, 0.041] | 0.0018 |
| C40-7 PC plasmalogen | 0.023 [0.0096, 0.037] | 0.0014 |
| C34-4 PC | 0.022 [0.0056, 0.038] | 0.011 |
| C38-6 PC plasmalogen | 0.022 [0.0084, 0.035] | 0.0022 |
| C36-5 PC plasmalogen-B | 0.019 [0.0059, 0.033] | 0.0066 |
| C38-4 PC plasmalogen | 0.017 [0.0063, 0.028] | 0.0031 |
| C36-4 PC-B | 0.017 [0.003, 0.03] | 0.02 |
| C38-4 PC | 0.016 [0.0037, 0.029] | 0.014 |
| C38-3 PC | 0.016 [0.0017, 0.03] | 0.033 |
| C18-1 LPC plasmalogen | -0.024 [-0.04, -0.0073] | 0.0065 |
| C16-1 LPC plasmalogen | -0.024 [-0.041, -0.0064] | 0.0095 |
| **Amino acids, peptides, and analogues** | | |
| Methionine | 0.042 [0.02, 0.063] | $4.5 \times 10^{-4}$ |
| Glutamine | 0.035 [0.01, 0.059] | 0.007 |
| N-acetylmethionine | 0.031 [0.013, 0.05] | 0.0013 |
| Asparagine | 0.028 [0.012, 0.044] | $9.1 \times 10^{-4}$ |
| Ornithine | 0.016 [0.00076, 0.032] | 0.045 |
| NMMA | 0.015 [0.0049, 0.025] | 0.0055 |
| N-acetyalanine | 0.012 [0.0035, 0.021] | 0.0082 |
| ADMA | 0.011 [0.00096, 0.022] | 0.037 |
| SDMA | 0.011 [0.0017, 0.02] | 0.024 |
| Creatinine | 0.0076 [0.002, 0.013] | 0.011 |
| Glutamate | -0.016 [-0.029, -0.0027] | 0.022 |
| Aspartate | -0.034 [-0.049, -0.02] | $2.6 \times 10^{-5}$ |
| Methionine sulfoxide | -0.048 [-0.067, -0.029] | $7.3 \times 10^{-6}$ |
| **Triacylglycerols** | | |
| C58-10 TAG +NH4 | 0.046 [0.0098, 0.083] | 0.016 |
| C58-6 TAG | 0.039 [0.015, 0.063] | 0.0024 |
| NH4_C44-1 TAG | 0.037 [0.0075, 0.067] | 0.017 |
| NH4_C46-3 TAG | 0.036 [0.0097, 0.063] | 0.0098 |
| NH4_C44-2 TAG | 0.036 [0.0068, 0.065] | 0.019 |
| C52-7 TAG | 0.035 [0.0064, 0.063] | 0.02 |
| C58-9 TAG +NH4 | 0.034 [0.0011, 0.066] | 0.048 |
| NH4_C46-2 TAG | 0.031 [0.0055, 0.057] | 0.021 |
| C56-8 TAG +NH4 | 0.031 [0.0041, 0.057] | 0.028 |
| C50-6 TAG | 0.03 [0.0021, 0.057] | 0.04 |
| C56-7 TAG +NH4 | 0.028 [0.006, 0.05] | 0.015 |
| C56-6 TAG +NH4 | 0.026 [0.0055, 0.046] | 0.015 |
| **Glycerophosphoethanolamines** | | |
| C38-5 PE | 0.03 [0.014, 0.046] | $6 \times 10^{-4}$ |
| C40-7 PE plasmalogen | 0.025 [0.01, 0.039] | 0.0012 |
| C40-6 PE | 0.019 [0.00082, 0.038] | 0.045 |
| C38-6 PE plasmalogen_2 | 0.018 [0.0054, 0.031] | 0.0072 |
| C36-4 PE plasmalogen | 0.017 [0.00089, 0.034] | 0.044 |
| C38-7 PE plasmalogen | 0.016 [0.002, 0.03] | 0.03 |
| C36-5 PE plasmalogen | 0.016 [0.0012, 0.031] | 0.039 |
| C38-6 PE | 0.016 [0.00079, 0.031] | 0.044 |
| C38-5 PE plasmalogen_2 | 0.015 [9e-04, 0.029] | 0.042 |
| C20-4 LPE | -0.019 [-0.031, -0.0063] | 0.0045 |
| **Diradylglycerols** | | |
| NH4_C34-1 DAG | 0.049 [0.024, 0.074] | $3.7 \times 10^{-4}$ |
| C38-4 DAG | 0.038 [0.019, 0.057] | $2.8 \times 10^{-4}$ |
| NH4_C34-3 DAG | 0.035 [0.0053, 0.064] | 0.025 |
| NH4_C36-2 DAG | 0.035 [0.016, 0.053] | $6 \times 10^{-4}$ |
| C38-5 DAG | 0.027 [0.0074, 0.047] | 0.0093 |
| NH4_C36-3 DAG | 0.026 [0.0078, 0.044] | 0.0068 |
| NH4_C34-2 DAG | 0.022 [0.00051, 0.042] | 0.049 |
| C36-0 DAG +NH4 | 0.013 [0.00043, 0.025] | 0.047 |
| C34-1 DAG | -0.039 [-0.057, -0.021] | $6.6 \times 10^{-5}$ |
| **Fatty acids and conjugates** | | |
| Butyrobetaine | 0.0095 [0.00065, 0.018] | 0.04 |
| Arachidonate | -0.017 [-0.029, -0.0057] | 0.0053 |
| 3-methyladipate | -0.031 [-0.06, -0.0018] | 0.042 |
| Azelate | -0.035 [-0.068, -0.0018] | 0.043 |
| 10-HDoHE_14-HDoHE | -0.04 [-0.06, -0.019] | $3.6 \times 10^{-4}$ |
| 11-HDoHE | -0.047 [-0.067, -0.027] | $3 \times 10^{-5}$ |
| 7-HDoHE_8-HDoHE | -0.049 [-0.072, -0.026] | $8.5 \times 10^{-5}$ |
| 16-HDoHE_17-HDoHE | -0.049 [-0.07, -0.029] | $1.4 \times 10^{-5}$ |
| 20-HDoHE | -0.054 [-0.077, -0.031] | $3.3 \times 10^{-5}$ |
| **Benzoic acids and derivatives** | | |
| 4-hydroxyhippurate | 0.031 [0.01, 0.051] | 0.0049 |
| Salicylurate | 0.03 [0.0096, 0.05] | 0.0053 |
| Methyl N-methylanthranilate | 0.03 [0.009, 0.05] | 0.0067 |
| Salicylate | 0.029 [0.012, 0.047] | 0.002 |
| 3-hydroxybenzoate | 0.017 [0.0034, 0.032] | 0.018 |
| Anthranilic acid | -0.031 [-0.05, -0.011] | 0.0031 |
| **Steroid esters** | | |
| C22-6 CE | 0.034 [0.016, 0.052] | $4.9 \times 10^{-4}$ |
| C20-5 CE | 0.029 [0.011, 0.047] | 0.0022 |
| C20-3 CE | 0.019 [0.0058, 0.033] | 0.0068 |
| C20-4 CE | 0.018 [0.0049, 0.03] | 0.0084 |
| **Fatty acid esters** | | |
| C2 carnitine | 0.031 [0.013, 0.049] | 0.0016 |
| C16 carnitine | 0.022 [0.0049, 0.038] | 0.014 |
| C3 carnitine | 0.018 [0.0034, 0.033] | 0.019 |
| **Sphingolipids** | | |
| Sphinganine | 0.031 [0.0062, 0.055] | 0.017 |
| Sphingosine | 0.029 [0.0046, 0.054] | 0.024 |
| L-threo-sphingosine | 0.027 [0.0024, 0.052] | 0.036 |
| **Tricarboxylic acids and derivatives** | | |
| Isocitrate | 0.018 [0.0024, 0.034] | 0.028 |
| Aconitate | 0.013 [0.00058, 0.026] | 0.045 |
| Citrate | 0.011 [0.0019, 0.02] | 0.02 |
| **Linoleic acids and derivatives** | | |
| Linoleate | -0.015 [-0.029, -0.0014] | 0.035 |
| 13-HODE | -0.056 [-0.076, -0.035] | $1.3 \times 10^{-6}$ |
| 13-HpODE | -0.062 [-0.083, -0.04] | $5.7 \times 10^{-7}$ |
| **Carbonyl compounds** | | |
| 4-hydroxybenzaldehyde | -0.016 [-0.032, -0.00042] | 0.049 |
| 4-hydroxy-3-methylacetophenone | -0.048 [-0.068, -0.028] | $2.1 \times 10^{-5}$ |
| **Gamma-keto acids and derivatives** | | |
| Levulinate | 0.069 [0.041, 0.097] | $9.6 \times 10^{-6}$ |
| **Sulfinic acids** | | |
| Hypotaurine | 0.048 [0.023, 0.074] | $4.5 \times 10^{-4}$ |
| **Hydroxyindoles** | | |
| 5-Hydroxytryptophol | 0.038 [0.0093, 0.067] | 0.012 |
| **Glycerophosphoserines** | | |
| C34-0 PS | 0.027 [0.0057, 0.048] | 0.016 |
| **Amines** | | |
| Spermidine | 0.02 [0.00099, 0.039] | 0.044 |
| **Dicarboxylic acids and derivatives** | | |
| Fumarate_maleate | 0.016 [0.0027, 0.029] | 0.021 |
| **Beta hydroxy acids and derivatives** | | |
| Malate | 0.015 [0.0019, 0.028] | 0.029 |
| **Purines and purine derivatives** | | |
| 7-methylguanine | 0.012 [0.00033, 0.023] | 0.048 |

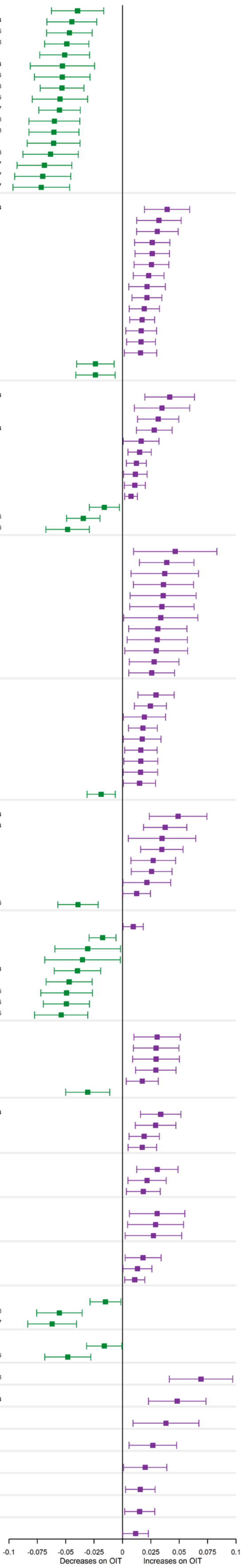


A.

| Metabolites | OR [95% CI] | p-value (GLM) |
|---|---|---|
| **Eicosanoids** | | |
| 5,6 DHET | -0.015 [-0.033, 0.002] | 0.091 |
| 12-HEPE | -0.04 [-0.063, -0.017] | 0.0014 |
| 15-HEPE | -0.045 [-0.066, -0.023] | $1.9 \times 10^{-4}$ |
| 12-HETE_8-HETE | -0.047 [-0.067, -0.027] | $2.8 \times 10^{-5}$ |
| 15-HETE | -0.049 [-0.069, -0.03] | $7.7 \times 10^{-6}$ |
| 11-HEPE | -0.051 [-0.073, -0.029] | $3 \times 10^{-5}$ |
| 5-HEPE | -0.053 [-0.081, -0.025] | $5.5 \times 10^{-4}$ |
| 9-HEPE | -0.053 [-0.078, -0.029] | $8.4 \times 10^{-5}$ |
| 11-HETE | -0.053 [-0.072, -0.034] | $1.3 \times 10^{-6}$ |
| LTB3 | -0.055 [-0.079, -0.031] | $4.5 \times 10^{-5}$ |
| 9-HETE | -0.055 [-0.074, -0.037] | $1.9 \times 10^{-7}$ |
| 5-HETE | -0.06 [-0.082, -0.038] | $2.4 \times 10^{-6}$ |
| Lipoxin A4 | -0.06 [-0.082, -0.038] | $1.6 \times 10^{-6}$ |
| 8,15 diHETE | -0.061 [-0.084, -0.037] | $4 \times 10^{-6}$ |
| 5,15 diHETE | -0.063 [-0.088, -0.039] | $4.4 \times 10^{-6}$ |
| 17,18 diHETE | -0.069 [-0.093, -0.045] | $7.1 \times 10^{-7}$ |
| 14,15 diHETE | -0.07 [-0.095, -0.046] | $7.1 \times 10^{-7}$ |
| LTB4 | -0.071 [-0.096, -0.046] | $6.5 \times 10^{-7}$ |
| **Bile acids, alcohols and derivatives** | | |
| Alpha-muricholate | 0.014 [-0.01, 0.038] | 0.27 |
| Lithocholate | 0.003 [-0.023, 0.029] | 0.83 |
| Glycolithocholate | 0.001 [-0.022, 0.024] | 0.95 |
| Chenodeoxycholate | -0.005 [-0.028, 0.018] | 0.69 |
| Hyodeoxycholate_ursodeoxycholate | -0.006 [-0.026, 0.013] | 0.52 |
| Glycochenodeoxycholate | -0.007 [-0.027, 0.012] | 0.46 |
| Glycodeoxycholate_glycochenodeoxycholate | -0.009 [-0.027, 0.01] | 0.38 |
| Taurochenodeoxycholate | -0.009 [-0.032, 0.013] | 0.42 |
| Glycocholate | -0.011 [-0.03, 0.009] | 0.29 |
| Glycoursodeoxycholate | -0.011 [-0.032, 0.01] | 0.3 |
| Taurocholate | -0.012 [-0.036, 0.012] | 0.33 |
| Cholate | -0.013 [-0.035, 0.009] | 0.25 |
| Deoxycholate | -0.013 [-0.035, 0.009] | 0.25 |
| Taurodeoxycholate_taurochenodeoxycholate | -0.013 [-0.036, 0.009] | 0.25 |
| Ketodeoxycholate | -0.015 [-0.041, 0.011] | 0.25 |
| Taurohyodeoxycholate_tauroursodeoxycholate | -0.016 [-0.04, 0.007] | 0.18 |
| Tauro-alpha-muricholate_tauro-beta-muricholate | -0.025 [-0.05, 0.001] | 0.068 |
| **Sphingolipids** | | |
| Sphinganine | 0.031 [0.006, 0.055] | 0.017 |
| Sphingosine | 0.029 [0.005, 0.054] | 0.024 |
| L-threo-sphingosine | 0.027 [0.002, 0.052] | 0.036 |
| C22-0 Ceramide (d18-1) | 0.008 [-0.006, 0.023] | 0.26 |
| C24-0 Ceramide (d18-1) | 0.006 [-0.006, 0.019] | 0.3 |
| C22-0 SM | 0.005 [-0.007, 0.017] | 0.42 |
| C24-1 Ceramide (d18-1) | 0.005 [-0.008, 0.017] | 0.47 |
| C14-0 SM | 0.004 [-0.009, 0.018] | 0.52 |
| C24-0 SM | 0.004 [-0.007, 0.016] | 0.48 |
| C16-0 Ceramide (d18-1) | 0.002 [-0.01, 0.014] | 0.74 |
| C16-0 SM | 0 [-0.011, 0.01] | 0.95 |
| C22-1 SM | -0.001 [-0.012, 0.011] | 0.92 |
| C24-1 SM | -0.002 [-0.012, 0.008] | 0.7 |
| C18-1 SM | -0.002 [-0.014, 0.01] | 0.74 |
| C20-0 SM | -0.002 [-0.015, 0.011] | 0.71 |
| C18-0 SM | -0.004 [-0.016, 0.008] | 0.52 |
| **Amino acids (histidine metabolism)** | | |
| Urocanic acid | 0.01 [-0.005, 0.025] | 0.2 |
| Histidine | 0.005 [-0.008, 0.017] | 0.47 |
| 1-methylhistidine | 0.003 [-0.011, 0.017] | 0.67 |
| Imidazole propionate | 0.001 [-0.017, 0.02] | 0.9 |
| 3-methylhistidine | -0.002 [-0.024, 0.021] | 0.89 |
| Methylimidazole acetic acid | -0.009 [-0.029, 0.01] | 0.35 |
| Carnosine | -0.027 [-0.058, 0.003] | 0.087 |
| **Linoleic acids and derivatives** | | |
| C36-3 DAG | 0.002 [-0.015, 0.019] | 0.81 |
| Gamma-linolenate | -0.005 [-0.023, 0.012] | 0.55 |
| Linoleate | -0.015 [-0.029, -0.001] | 0.035 |
| Alpha-linolenate | -0.016 [-0.032, 0.001] | 0.073 |
| 13-HODE | -0.056 [-0.076, -0.035] | $1.3 \times 10^{-6}$ |
| 13-HpODE | -0.062 [-0.083, -0.04] | $5.7 \times 10^{-7}$ |

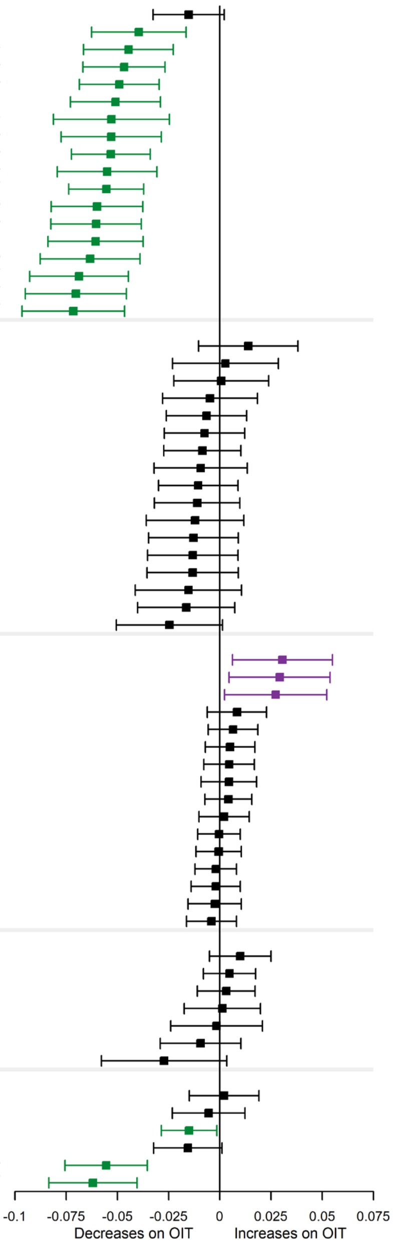


**B.**

**Supplemental Figure E2 – PNOIT: Metabolites associated with OIT outcome.** Generalized linear model of metabolites significantly associated OIT outcome (higher in SU in blue, higher in TD in red) adjusted for age and time on OIT, with (A) all significant metabolites, and (B) all identified metabolites in the key pathways of interest (bile acids, histidine metabolites, sphingolipids, eicosanoid & linoleic acid metabolites).

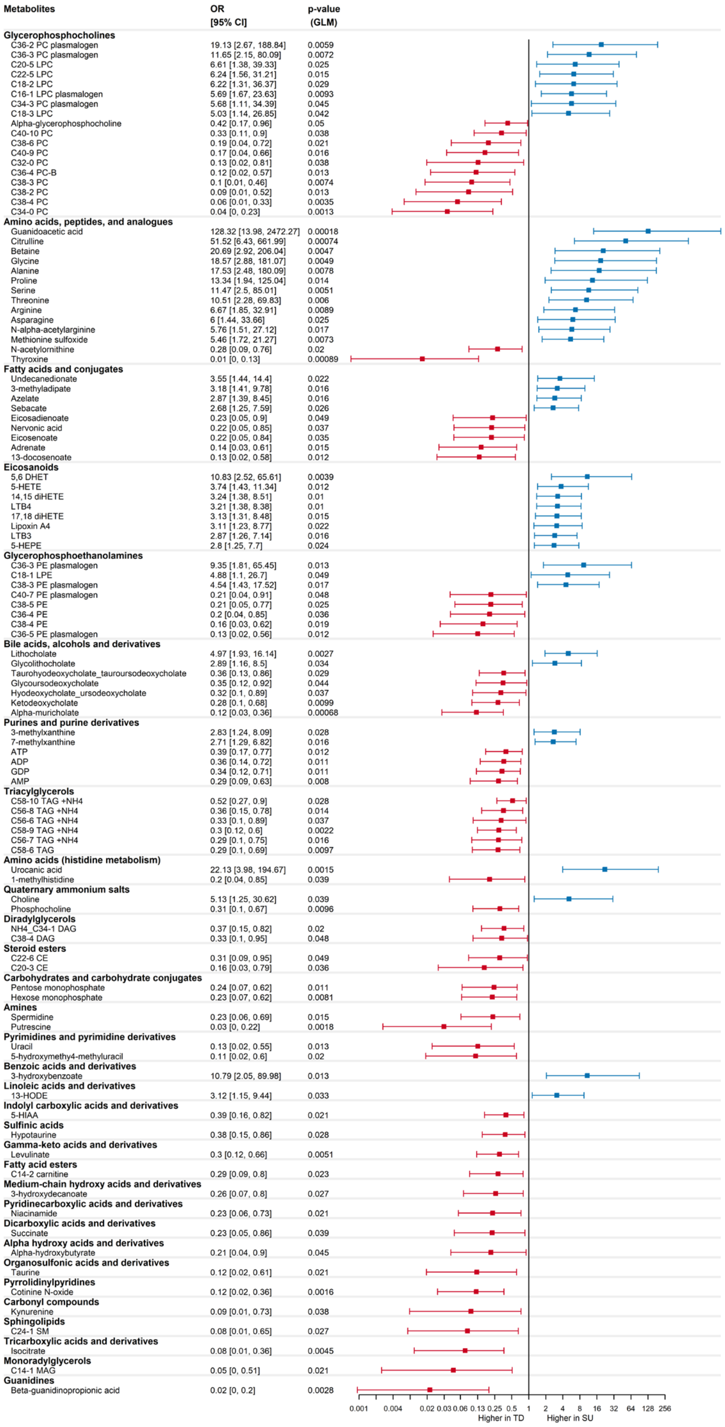


A.

| Metabolites | OR [95% CI] | p-value (GLM) |
|---|---|---|
| **Eicosanoids** | | |
| 5,6 DHET | 10.83 [2.52, 65.61] | 0.0039 |
| 5-HETE | 3.74 [1.43, 11.34] | 0.012 |
| 14,15 diHETE | 3.24 [1.38, 8.51] | 0.01 |
| LTB4 | 3.21 [1.38, 8.38] | 0.01 |
| 17,18 diHETE | 3.13 [1.31, 8.48] | 0.015 |
| Lipoxin A4 | 3.11 [1.23, 8.77] | 0.022 |
| LTB3 | 2.87 [1.26, 7.14] | 0.016 |
| 5-HEPE | 2.8 [1.25, 7.7] | 0.024 |
| 8,15 diHETE | 2.39 [1.02, 6.14] | 0.054 |
| 12-HETE_8-HETE | 2.2 [0.82, 6.69] | 0.13 |
| 5,15 diHETE | 2.14 [0.96, 5.16] | 0.073 |
| 15-HETE | 2.1 [0.77, 6.29] | 0.16 |
| 11-HEPE | 2.06 [0.84, 5.56] | 0.13 |
| 9-HETE | 1.98 [0.69, 6.17] | 0.22 |
| 11-HETE | 1.94 [0.71, 5.68] | 0.21 |
| 9-HEPE | 1.81 [0.81, 4.5] | 0.17 |
| 12-HEPE | 1.56 [0.67, 3.87] | 0.31 |
| 15-HEPE | 1.47 [0.61, 3.73] | 0.39 |
| **Bile acids, alcohols and derivatives** | | |
| Lithocholate | 4.97 [1.93, 16.14] | 0.0027 |
| Glycolithocholate | 2.89 [1.16, 8.5] | 0.034 |
| Glycochenodeoxycholate | 1.79 [0.67, 5.08] | 0.26 |
| Glycocholate | 1.74 [0.64, 5.13] | 0.29 |
| Glycodeoxycholate_glycochenodeoxycholate | 1.08 [0.39, 3.07] | 0.88 |
| Taurodeoxycholate_taurochenodeoxycholate | 0.95 [0.4, 2.25] | 0.9 |
| Taurochenodeoxycholate | 0.89 [0.38, 2.07] | 0.79 |
| Taurocholate | 0.89 [0.39, 2] | 0.78 |
| Tauro-alpha-muricholate_tauro-beta-muricholate | 0.83 [0.39, 1.77] | 0.63 |
| Cholate | 0.79 [0.31, 1.9] | 0.6 |
| Chenodeoxycholate | 0.55 [0.22, 1.28] | 0.18 |
| Deoxycholate | 0.51 [0.19, 1.22] | 0.14 |
| Taurohyodeoxycholate_tauroursodeoxycholate | 0.36 [0.13, 0.86] | 0.029 |
| Glycoursodeoxycholate | 0.35 [0.12, 0.92] | 0.044 |
| Hyodeoxycholate_ursodeoxycholate | 0.32 [0.1, 0.89] | 0.037 |
| Ketodeoxycholate | 0.28 [0.1, 0.68] | 0.0099 |
| Alpha-muricholate | 0.12 [0.03, 0.36] | 0.00068 |
| **Sphingolipids** | | |
| C16-0 SM | 2.8 [0.43, 21.67] | 0.29 |
| C24-1 Ceramide (d18-1) | 1.33 [0.28, 6.64] | 0.72 |
| C18-1 SM | 1.21 [0.24, 6.22] | 0.81 |
| C14-0 SM | 1.12 [0.27, 4.74] | 0.88 |
| C18-0 SM | 0.99 [0.2, 4.87] | 0.99 |
| C20-0 SM | 0.98 [0.22, 4.42] | 0.98 |
| C24-0 Ceramide (d18-1) | 0.89 [0.18, 4.38] | 0.88 |
| C22-0 Ceramide (d18-1) | 0.75 [0.19, 2.82] | 0.67 |
| Sphinganine | 0.6 [0.25, 1.32] | 0.21 |
| C22-1 SM | 0.56 [0.09, 3.17] | 0.52 |
| Sphingosine | 0.54 [0.22, 1.2] | 0.14 |
| C16-0 Ceramide (d18-1) | 0.43 [0.08, 2.07] | 0.31 |
| C24-0 SM | 0.29 [0.05, 1.59] | 0.17 |
| C22-0 SM | 0.18 [0.03, 0.98] | 0.061 |
| C24-1 SM | 0.08 [0.01, 0.65] | 0.027 |
| **Amino acids (histidine metabolism)** | | |
| Urocanic acid | 22.13 [3.98, 194.67] | 0.0015 |
| Histidine | 5.08 [0.96, 40.38] | 0.082 |
| Carnosine | 1.31 [0.7, 2.54] | 0.41 |
| Imidazole propionate | 0.79 [0.28, 2.22] | 0.66 |
| Methylimidazole acetic acid | 0.45 [0.16, 1.22] | 0.13 |
| 3-methylhistidine | 0.43 [0.16, 1.04] | 0.074 |
| 1-methylhistidine | 0.2 [0.04, 0.85] | 0.039 |
| **Linoleic acids and derivatives** | | |
| 13-HODE | 3.12 [1.15, 9.44] | 0.033 |
| 13-HpODE | 2.08 [0.84, 5.61] | 0.13 |
| Alpha-linolenate | 0.71 [0.22, 2.25] | 0.56 |
| Linoleate | 0.58 [0.13, 2.4] | 0.46 |
| C36-3 DAG | 0.57 [0.17, 1.76] | 0.33 |
| Gamma-linolenate | 0.53 [0.16, 1.6] | 0.28 |

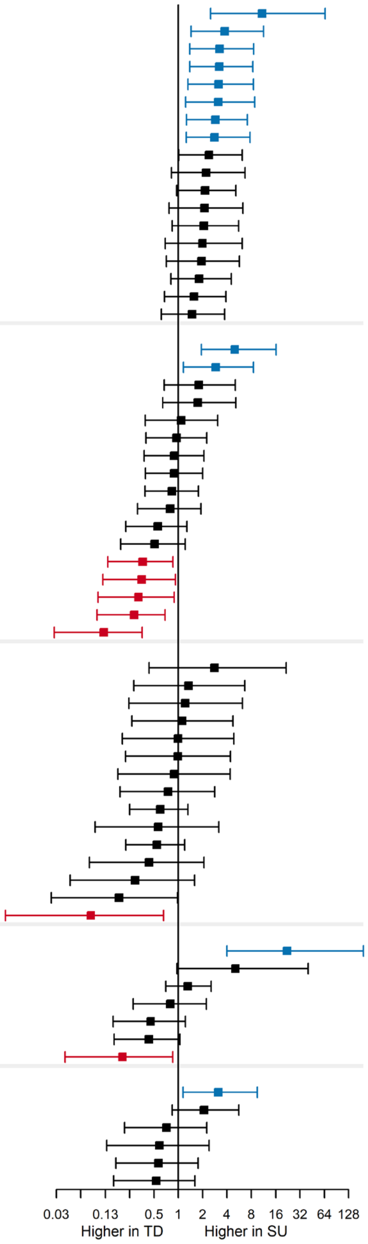


B.

**Supplemental Table E1: PNOIT: Biological pathways associated with time on OIT matched to metabolites of interest**.

| Pathway Name | Pathway Source/Pathway ID | p-value (FDR adj) (% Matched Analytes) | Matched Analytes |
|---|---|---|---|
| Metabolism of alpha-linolenic acid | wiki \| WP4586 | 0.000011 (35%) | Leinolic acid; Linoleic acid; all-cis-5,8,11,14-Eicosatetraenoic acid; Arachidonic acid;15-HETE; FA 20:4;O;5S,6R-LipoxinA4; Lipoxin A4;12-HEPE; FA 20:5;O;15-HEPE; FA 20:5;O;5-HETE |
| Eicosanoid synthesis | wiki \| WP167 | 0.02 (15%) | all-cis-5,8,11,14-Eicosatetraenoic acid; Arachidonic acid;15-HETE; FA 20:4;O;11(R)-HETE;5-HETE |
| Transport of inorganic cations/anions and amino acids | wiki \| WP1936 | 0.00016 (15%) | Glutamate; L-Glutamic acid;(S)-malate; Malic acid;L-Asparagine;L-Aspartic acid;ornithine;Glutamine; L-Glutamine;L-Methionine; Methionine;L-Targinine; methylArg |
| Sphingolipid metabolism in senescence | wiki \| WP5121 | 0.025 (21%) | Sphingosine;D-erythro-2-Amino-1,3-octadecanediol; Sphinganine;DG(16:0_16:0); DG(18:0/20:4(5Z,8Z,11Z,14Z)/0:0) |
| Alanine and aspartate metabolism | wiki \| WP106 | 0.0001 (29%) | Citric acid;fumarate; Fumaric acid;(S)-malate; Malic acid;L-Asparagine;L-Aspartic acid;L-Acetylcarnitine; O-Acetylcarnitine |
| Eicosanoid metabolism via lipooxygenases (LOX) | wiki \| WP4721 | 0.00072 (19%) | all-cis-5,8,11,14-Eicosatetraenoic acid; Arachidonic acid;15-HETE; FA 20:4;O;5S,6R-LipoxinA4; Lipoxin A4;8-HETE;12(S)-Leukotriene B4; 6t,12epi-LTB4;5-HETE |

| | | | |
|---|---|---|---|
| Metabolism overview | wiki \| WP3602 | 0.00091 (9%) | Citric acid;fumarate; Fumaric acid;Glutamate; L-Glutamic acid;(S)-malate; Malic acid;L-Aspartic acid;D-threo-Isocitrate; Isocitric acid;Ornithine;L-Palmitoylcarnitine; L-PCARN; Palmitoylcarnitine;Glutamine; L-Glutamine; TG(16:0/20:4(5Z,8Z,11Z,14Z)/20:4(5Z,8Z,11Z,14Z)); TG(18:1(9Z)/18:1(9Z)/20:4(5Z,8Z,11Z,14Z)); TG(18:1(9Z)/20:1(11Z)/20:4(5Z,8Z,11Z,14Z)); TG(18:1(9Z)/18:2(9Z,12Z)/20:4(5Z,8Z,11Z,14Z)); TG(18:1(9Z)/20:4(5Z,8Z,11Z,14Z)/20:4(5Z,8Z,11Z,14Z)); TG(18:2(9Z,12Z)/20:4(5Z,8Z,11Z,14Z)/20:4(5Z,8Z,11Z,14Z)) |
| Urea cycle and related diseases | wiki \| WP4571 | 0.00016 (36%) | fumarate; Fumaric acid;Glutamate; L-Glutamic acid;L-Aspartic acid;ornithine;Glutamine; L-Glutamine |
| Metabolic reprogramming in colon cancer | wiki \| WP4290 | 0.00016 (18%) | Citric acid;fumarate; Fumaric acid;Glutamate; L-Glutamic acid;(S)-malate; Malic acid;L-Asparagine;D-threo-Isocitrate; Isocitric acid;Glutamine; L-Glutamine |
| Urea cycle and metabolism of amino groups | wiki \| WP497 | 0.00091 (18%) | fumarate; Fumaric acid;Glutamate; L-Glutamic acid;L-Aspartic acid;ornithine;Creatinine;Spermidine |
| Arachidonic acid (AA, ARA) oxylipin metabolism | wiki \| WP5155 | 0.000076 (13%) | all-cis-5,8,11,14-Eicosatetraenoic acid; Arachidonic acid;15-HETE; FA 20:4;O;5S,6R-LipoxinA4; Lipoxin A4;8-HETE;11(R)-HETE;12(S)-Leukotriene B4; 6t,12epi-LTB4;5,15-DiHETE; FA 20:4;O2;8,15-DiHETE; FA 20:4;O2;9-HETE;5-HETE |
| Biomarkers for urea cycle disorders | wiki \| WP4583 | 0.00011 (27%) | fumarate; Fumaric acid;Glutamate; L-Glutamic acid;L-Aspartic acid;ornithine;Glutamine; L-Glutamine;L-Methionine; Methionine |

| | | | |
|---|---|---|---|
| Amino acid metabolism | wiki \| WP3925 | 0.00004 (11%) | cis-aconitate; cis-Aconitic acid;Citric acid;fumarate; Fumaric acid;Glutamate; L-Glutamic acid;(S)-malate; Malic acid;L-Asparagine;L-Aspartic acid;D-threo-Isocitrate; Isocitric acid;Ornithine;Glutamine; L-Glutamine;L-Methionine; Methionine;Spermidine |
| Hereditary leiomyomatosis and renal cell carcinoma pathway | wiki \| WP4206 | 0.00091 (24%) | Citric acid;fumarate; Fumaric acid;Glutamate; L-Glutamic acid;(S)-malate; Malic acid;D-threo-Isocitrate; Isocitric acid |
| Amino acid metabolism pathway excerpt: histidine catabolism extension | wiki \| WP4661 | 0.00004 (35%) | cis-aconitate; cis-Aconitic acid;Citric acid;fumarate; Fumaric acid;Glutamate; L-Glutamic acid;(S)-malate; Malic acid;D-threo-Isocitrate; Isocitric acid |
| Transport of bile salts and organic acids, metal ions and amine compounds | reactome \| R-HSA-425366 | 0.05 (9%) | Citric acid;L-Asparagine;Creatinine;Glutamine; L-Glutamine;L-Methionine; Methionine;Spermidine;SAL; Salicylic acid |

**Supplemental Table E2: PNOIT: Biological pathways associated with OIT outcome matched to metabolites of interest**.

| Pathway Name | Pathway Source/Pathway ID | p-value (FDR adj) (% Matched Analytes) | Matched Analytes |
|---|---|---|---|
| Primary bile acid biosynthesis | kegg \| map00120 | 0.017 (17%) | Glycine;Taurine;Adenosine triphosphate; ATP;Lithocholic acid glycine conjugate;Glycoursodeoxycholate; Glycoursodeoxycholic acid;Lithocholic acid; ST 24:1;O3;Tauroursodeoxycholic acid;Ursodeoxycholate; Ursodeoxycholic acid;ADP;CE(22:6(4Z,7Z,10Z,13Z,16Z,19Z));CE(20:3(8Z,11Z,14Z)) |

| | | | |
|---|---|---|---|
| Transport of bile salts and organic acids, metal ions and amine compounds | reactome \| R-HSA-425366 | 0.000054 (15%) | Betaine;Choline;Glycine;L-Alanine;L-Proline;L-Threonine;L-Asparagine;L-Serine; Serine;Taurine;succinate; Succinic acid; Wormwood acid;Arginine; L-Arginine;Spermidine |
| Recycling of bile acids and salts | reactome \| R-HSA-159418 | 0.0000002 (45%) | Adenosine monophosphate; AMP;Glycine;Taurine;Adenosine triphosphate; ATP;Glycoursodeoxycholate; Glycoursodeoxycholic acid;Lithocholic acid; ST 24:1;O3;Tauroursodeoxycholic acid;Ursodeoxycholate; Ursodeoxycholic acid;ADP |
| Synthesis of bile acids and bile salts | reactome \| R-HSA-193368 | 0.00019 (18%) | Adenosine monophosphate; AMP;Glycine;Taurine;Adenosine triphosphate; ATP;Glycoursodeoxycholate; Glycoursodeoxycholic acid;Lithocholic acid; ST 24:1;O3;Tauroursodeoxycholic acid;Ursodeoxycholate; Ursodeoxycholic acid;ADP |
| Histidine catabolism | wiki \| WP4988 | 0.015 (14%) | Urocanic acid;Adenosine triphosphate; ATP;ADP |
| Sphingolipid metabolism | wiki \| WP2788 | 0.0037 (9%) | L-Serine; Serine;Adenosine triphosphate; ATP;ADP;Phosphorylcholine;DG(16:0_16:0); DG(18:0/20:4(5Z,8Z,11Z,14Z)/0:0);ceramide phosphocholine (sphingomyelin, SM); N-lauroyl-D-erythro-Sphingosylphosphoryl choline; SM(d18:1/24:1(15Z)) |
| Eicosanoid metabolism via lipooxygenases (LOX) | wiki \| WP4721 | 0.03 (9%) | 5S,6R-LipoxinA4; Lipoxin A4;12(S)-Leukotriene B4; 6t,12epi-LTB4;5-HETE |
